# Synergic activity of FGFR2 and MEK inhibitors in the treatment of FGFR2-amplified cancers of unknown primary

**DOI:** 10.1101/2023.03.12.23287041

**Authors:** Andrea Cavazzoni, Irene Salamon, Claudia Fumarola, Giulia Gallerani, Noemi Laprovitera, Francesco Gelsomino, Mattia Riefolo, Karim Rihawi, Elisa Porcellini, Tania Rossi, Martina Mazzeschi, Maria Naddeo, Salvatore Serravalle, Elisabetta Broseghini, Federico Agostinis, Olivier Deas, Roberta Roncarati, Giorgio Durante, Mattia Lauriola, Ingrid Garajova, George A. Calin, Massimiliano Bonafè, Antonia D’Errico, Pier Giorgio Petronini, Stefano Cairo, Andrea Ardizzoni, Gabriele Sales, Manuela Ferracin

## Abstract

Patients with cancer of unknown primary (CUP) carry the burden of an aggressive disease and reduced access to therapies. Experimental models are pivotal for CUP biology investigation and drug testing. We derived two CUP cell lines (CUP#55 and #96), and corresponding patient-derived xenografts (PDXs), from ascites tumor cells. CUP cell lines and PDXs underwent histological, immune-phenotypical, molecular, and genomic characterization confirming the features of the original tumor. The tissue-of-origin prediction was obtained from the tumor microRNA expression profile and confirmed by single-cell transcriptomics. Genomic testing and FISH analysis identified FGFR2 gene amplification in both models, in the form of homogenously staining region (HSR) in CUP#55 and double minutes in CUP#96. FGFR2 was recognized as the main oncogenic driver and therapeutic target. FGFR2-targeting drug BGJ-398 (infigratinib) in combination with the MEK inhibitor trametinib proved to be synergic and exceptionally active, both *in vitro* and *in vivo*. The effects of the combined treatment by single-cell gene expression analysis revealed a remarkable plasticity of tumor cells and the greater sensitivity of cells with epithelial phenotype. This study brings personalized therapy closer to CUP patients and provides the rationale for FGFR2 and MEK targeting in metastatic tumors with FGFR2 pathway activation.

## INTRODUCTION

Metastases develop when tumor cells spread from the primary site to surrounding or distant tissues and are responsible for 90% of cancer-related deaths (1). Among metastatic patients, 3-5% show no clinical evidence of a primary site at diagnosis. These cases are classified as cancers of uncertain origin or cancers of unknown primary site (CUPs) or occult primary tumors (2) (3). They are usually diagnosed at a late stage, with patients presenting one or more metastases already at first diagnosis. The identification of tumor primary site is usually obtained by a combination of diagnostic investigations including physical examinations, blood analyses, imaging and immunohistochemical (IHC) testing of the tumor tissue. In CUP patients, these investigations are inconclusive.

International guidelines for tumor treatment are based on primary site indication. Therefore, CUP treatment requires a blind approach, which is very challenging for oncologists. Consequently, CUPs are typically treated with empiric platinum-based chemotherapy regimens, which are usually poorly effective. Indeed, CUP patients have a short life-expectancy (average overall survival 4-9 months, with only 20% surviving more than 1 year) which, unfortunately, did not improve over the last decades. However, these regimens remain empirical since they are mostly based on results of single-arm phase II clinical trials (4) (5) (6) or small phase III trials (7) (8) (9).

The use of molecular tests based on gene/miRNA expression signatures or methylation profiles to identify the most probable site-of-origin could assist the oncologists in the selection of the best treatment options and potentially improve CUPs outcome (10) (11). NCCN guidelines for occult primary tumors recently introduced the recommendation to use NGS genetic testing to guide therapeutic decision (v.2/2023) and suggest 11 different chemotherapy regimens for adenocarcinoma and 9 for squamous histology.

With the advent of personalized medicine, patient management is more and more frequently associated with the identification of specific molecular or genetic features of the tumor upon which therapies could be based to avoid suboptimal treatments. The identification of druggable alterations in CUP tumors could increase the otherwise limited treatment options, as recently demonstrated by Hayashi et al. (12). Next generation sequencing technologies were applied to the analysis of CUP mutational profile (13) (14) (15) (16) (17). Overall, CUPs seem to harbor recognized actionable genetic alterations in nearly 30% of cases (3) (18). Immunotherapy has been scarcely tested on CUP patients: in a study by Gatalica et al. only a fraction of patients presented favorable biomarkers for the use of immune checkpoint inhibitors (19). Varghese et al. identified the actionable mutations in a dataset of 150 CUPs analyzed with the MSK-IMPACT panel and patients who were treated with targeted therapies, showed clinical benefit and longer survival (15). A meta-analysis conducted by Ding et al. confirmed a benefit for site-specific therapies only for CUP patients with a stronger primary site prediction (20).

However, the knowledge that molecular and genetic testing could provide novel personalized treatments for CUP patients is hampered by the lack of cellular and animal models on which to test potentially effective therapies.

A main limitation in the development of CUP models is posed by the reduced availability of fresh tumor cells, given that the biopsy is frequently entirely dedicated to the diagnostic workup and surgery is rarely an option for these patients. Circulating tumor cells have been tracked in the blood of CUP patients (10) and could be the source of tumor cells for cell lines development; this is true also for ascites fluid, when available. Liquid biopsy tumor cells would have the advantage of being more representative of the overall complexity and heterogeneity of CUP tumors.

Recently, Verginelli et al. described the generation of the first CUP *in vitro* and *in vivo* models from biopsy/surgery tumor tissue, showing how they recapitulate the genetic of the original tumors and present a stem cell-like phenotype (21). Since tumor cell proliferation in their models was sustained by constitutively activation of the MAPK pathway, they described how the use of MEK1/2 inhibitor, trametinib, strongly reduced cell viability and tumor volume in xenograft models. This is the first study to describe the activation of an oncogenic pathway shared among CUPs, which supported a specific therapeutic intervention.

In this study, we obtained and expanded two patient-derived CUP cell lines from ascites tumor cells, spontaneously growing as spheroids and organoids, and corresponding patient-derived xenografts (PDXs). We obtained the immunophenotypic, molecular and genomic characterization of the tumor and derived models, confirming the recapitulation of the original features. Finally, we performed a drug-screening assay to target actionable genes and identified a combination of drugs with promising antitumor activity, both *in vitro* and *in vivo*.

## RESULTS

### Establishment of two CUP models from ascites tumor cells: immunophenotypical, genetic and molecular characterization

We generated *in vitro* and *in vivo* CUP models to test tailored experimental pharmacological approaches. CUP patient #55 was diagnosed with multiple lymph node metastases of poorly differentiated adenocarcinoma. Immunohistochemistry (IHC) staining reported the positivity for keratin 7 (K7) and CDX2, weak positivity for keratin 20 (K20) and negativity for neuroendocrine markers (chromogranin, synaptophysin, CD56). CUP patient #96 presented with multiple metastases of poorly differentiated adenocarcinoma with peritoneal carcinosis and multiple sub- and supra-diaphragmatic lymph node metastases. The tumor IHC staining was positive for K7, K20, CDX2, EpCAM and negative for PAX8, p40, GATA3 and calretinin.

Two long-term (more than 10 passages (22)) cell lines were obtained isolating cells from ascites fluid of the patients #55 and #96. One month after seeding, cells growing in suspension were visible as spheroid-like structures (**Figure S1**). The growth curve of two models was monitored for 10 days using the Incucyte S3 live-cell analysis: 10000 cells of CUP#96 and CUP#55 were seeded and after few hours CUP#96 cells formed large tumoroids and CUP#55 cells organized as clusters, showing a doubling time of 5 days for both cell lines (**Figure S2 and S3**).

We generated two PDX models by injecting ascites tumor cells in the interscapular region of immunocompromised mice as described in the Methods section. The two models showed different growth rate, with CUP#96 PDX growing faster than CUP#55 PDX, and with CUP#55 PDX inducing a tumor-intrinsic mild cachexia in the mice (**Figure S4**).

Morphology and histology of the cell lines and PDXs recapitulated the features of the primary tumor (**Figure 1A-B**). To verify whether CUP tumor cells presented a stem-cell like phenotype, the two tumoroid cell lines were tested for CD44 and EPCAM immunoreactivity and the expression of stemness genes CD44, NANOG and OCT4/POU5F1 (**Figure 2**). The two cell lines express CD44 on their surface, which is more homogeneous for CUP#96 (**Figure 2A**), and both express stemness genes at high levels if compared to a panel of cancer cell lines (**Figure 2B**).

**Figure 1.**
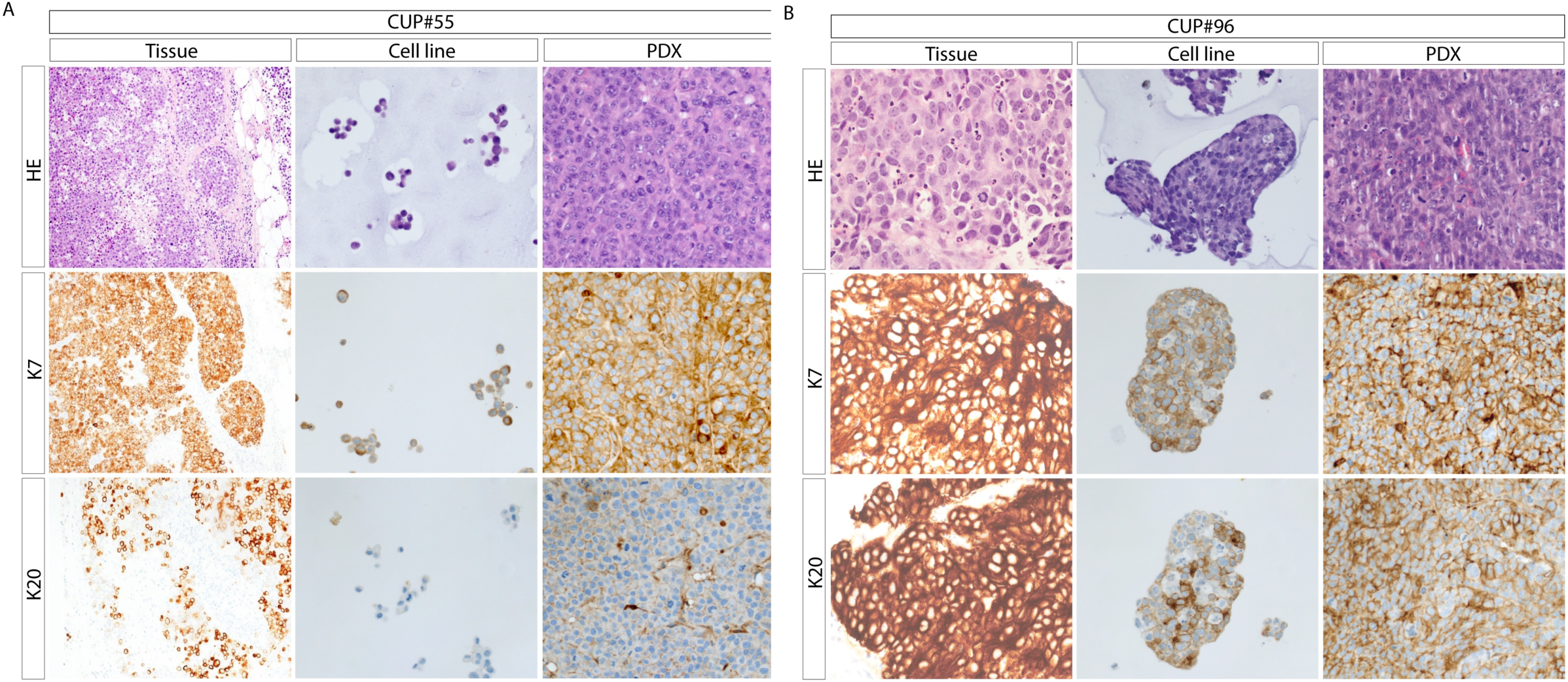
A) Immunophenotypic characterization of tumor tissues, cell lines and PDX of CUP#55. Hematoxylin-eosin (HE), keratin7 (K7) and keratin20 (K20) IHC staining was reported for tumor tissue, cell lines and PDX in CUP#55 model (40x). The staining showed that cell line and PDX models recapitulated the histology of the tumor tissue. **B) Immunophenotypic characterization of tumor tissues, cell lines and PDX of CUP#96.** Hematoxylin-eosin (HE), keratin7 (K7) and keratin20 (K20) IHC staining was reported for tumor tissue, cell lines and PDX in CUP#96 model (40x). CUP#96 cell line grows forming tumoroid structures. The staining showed that cell line and PDX models recapitulated the histology of the tumor tissue.

**Figure 2.**
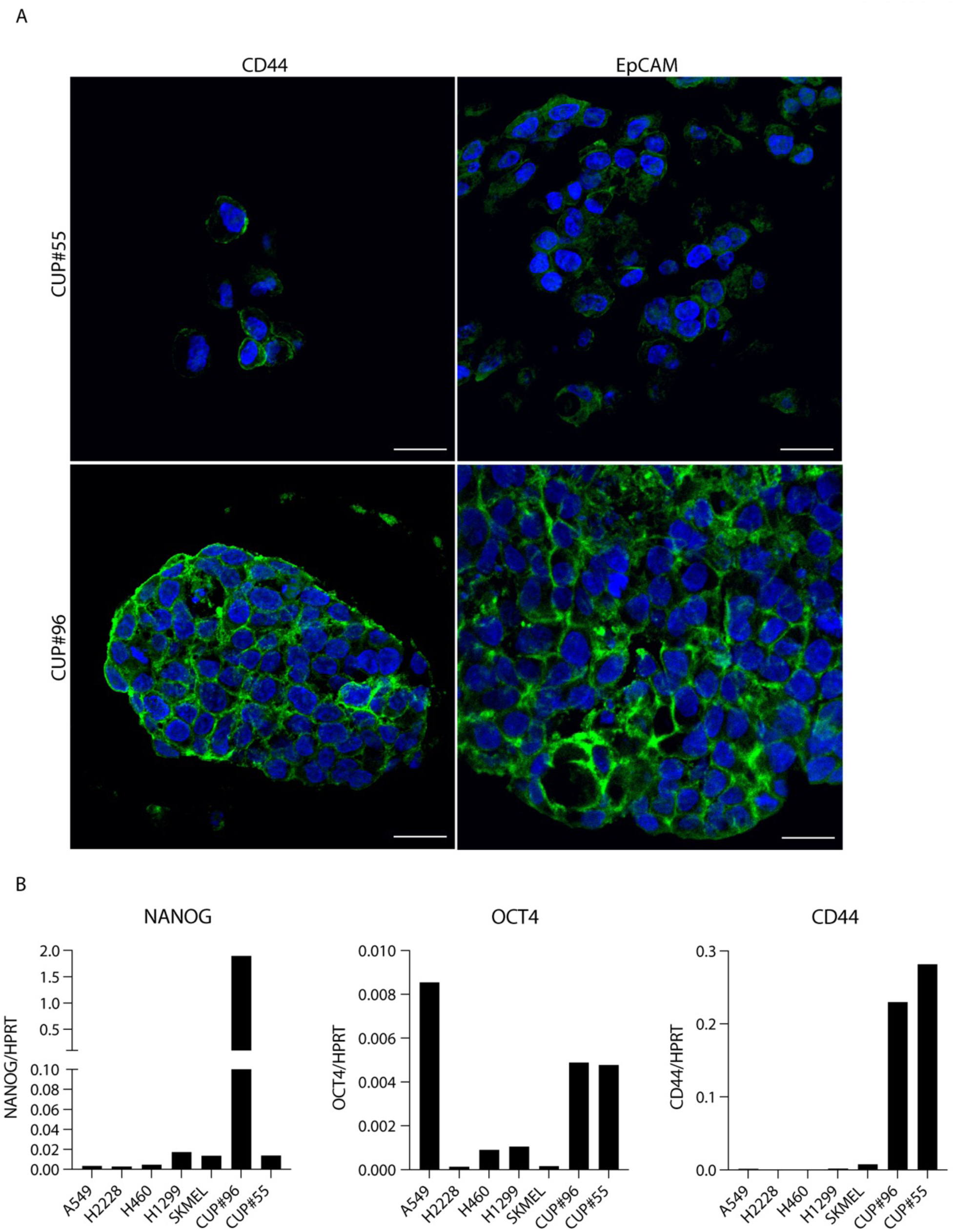
CUP models characterization. A) Immunophenotypic analysis of CUP #55 and #96 cell lines demonstrating the expression of cancer stem cell markers CD44 and EPCAM. Staining: nucleus (DAPI, blue), CD44 and EPCAM (green). B) Gene expression analysis of stemness genes in CUP#55 and CUP#96. Bars represent the ratio between the gene copies and the reference HPRT copies assessed by ddPCR. The expression in CUP cell lines was compared to other cancer cell lines from lung and melanoma origin.

We recently developed a predictive microRNA-based test to assign a possible primary site to metastatic cancers, including occult primary tumors (11). When we applied the predictive algorithms to the two tumors, we obtained a site-of-origin prediction: CUP#55 was predicted to be of biliary tract origin (probability of 93%) and CUP#96 of gastrointestinal tract origin (probability of 99%).

The tumor tissue, circulating cell-free DNA (ccfDNA), cell line and PDX were tested for genetic alterations with a CUP-dedicated, 92-gene custom panel using SureSelect Target Enrichment technology (Agilent Technologies) as described in (10). The summary of all genetic alterations is reported in **Table 1** and **Table 2**. The genetic analysis of CUP#55 confirmed the detection of 5 genetic alterations in all the analyzed samples (bulk tumor DNA, ccfDNA, cell line and PDX): the insertion in the *APC* gene (p.T1556fs*3) and the point mutations in *ARID1A* (p.R1276∗), *ERBB3* (p.S128R), *KEAP1* (p.R135H) and *NTRK1* (p.Q736X). Mutations in ALK, EPHA5, FAT1, KMT2C, MGA, PTPRD (except for p.L970V) and TP53 were detected only in ccfDNA (**Table 1**). As expected, the variant allele fractions (VAFs) were higher in PDX and cell line, thus reflecting the greater tumor purity and the possible selection of tumor subclones. Low frequency mutations in *ZFHX3, RBM10, PTPRT, NF1, MED12, MGA, KDR, KDM5A, GRIN2A, EP300, DOT1L, APC* (p. S887R), were identified in the models but not in other samples. The generic analysis of CUP#96 was conducted on ccfDNA and cell line due to the unavailability of residual tumor sample for this patient. Five genetic alterations were detected in both samples (**Table 2**), with higher VAFs in the cell line compared to ccfDNA. Low-pass Copy Number Analysis on the two cell lines revealed several regions of amplification and deletion, including a >10 copies amplification in chromosome 10 encompassing *FGFR2* oncogene (**Tables S1** and **S2**).

**Table 1.** Genetic alterations detected in CUP#55 patient-derived tissues and models.

| Gene | coding<br>change | aminoacidic<br>change | tumor FFPE | ccfDNA at<br>diagnosis | ccfDNA at<br>disease<br>progression | cell line | PDX Passage1 | predicted as<br>pathogenic* |
| --- | --- | --- | --- | --- | --- | --- | --- | --- |
| ALK | c.A1301G | p.K434R | not detected | not detected | 7.0% | not detected | not detected | no |
| APC | 4607insA | p.T1556fs*3 | 9.8% | 22.6% | 29.9% | 100.0% | 95.0% | ND |
| APC | c.A2659C | p.S887R | not detected | not detected | not detected | 43.2% | not detected | yes |
| ARID1A | c.C3826T | p.R1276X | 8.9% | 28.5% | 22.8% | 53.0% | 19.8% | yes |
| CREBBP | c.T1448C | p.L483P | not detected | not detected | not detected | not detected | 21.7% | ND |
| DOT1L | c.A1193C | p.K398T | not detected | not detected | not detected | 38.2% | not detected | yes |
| EP300 | c.A7118C | p.N2373T | not detected | not detected | not detected | not detected | 32.2% | no |
| EPHA5 | c.A1417C | p.T473P | not detected | not detected | 8.3% | not detected | not detected | no |
| ERBB3 | c.C384A | p.S128R | 9.8% | 20.7% | 20.4% | 41.09% | 48.60% | yes |
| ERBB4 | c.T343A | p.Y115N | not detected | not detected | not detected | not detected | 33.8% | yes |
| FAT1 | c.A2084C | p.N695T | not detected | not detected | 2.6% | not detected | not detected | no |
| GRIN2A | c.A4064C | p.K1355T | not detected | not detected | not detected | not detected | 22.3% | no |
| KDM5A | c.T3547C | p.W1183R | not detected | not detected | not detected | not detected | 22.2% | yes |
| KDR | c.A981C | p.K327N | not detected | not detected | not detected | not detected | 27.9% | ND |
| KEAP1 | c.G404A | p.R135H | 9.0% | 22.6% | 19.5% | 51.8% | 44.0% | yes |
| KMT2C | c.C2459T | p.T820I | not detected | 1.1% | 1.1% | not detected | not detected | yes |
| KMT2C | c.C1013T | p.S338L | not detected | 5.5% | 4.3% | not detected | not detected | no |
| KMT2C | c.C2689T | p.R897X | not detected | not detected | 1.4% | not detected | not detected | yes |
| KMT2C | c.C2228T | p.P743L | not detected | not detected | 1.6% | not detected | not detected | no |
| MED12 | c.A688C | p.I230L | not detected | not detected | not detected | not detected | 25.0% | no |
| MGA | c.A395C | p.N132T | not detected | not detected | not detected | not detected | 30.5% | yes |
| MGA | c.T4004C | p.L1335P | not detected | not detected | 1.6% | not detected | not detected | ND |
| NF1 | c.A6083G | p.K2028R | not detected | not detected | not detected | not detected | 25.4% | yes |
| NTRK1 | c.C2206T | p.Q736X | 8.3% | 26.0% | 22.3% | 32.6% | 44.7% | yes |
| PALB2 | C2419T | p.P807S | 9.0% | 19.1% | 16.6% | not detected | 47.4% | no |
| PTPRD | c.T2908G | p.L970V | not detected | 5.1% | 3.5% | not detected | 47.7% | yes |
| PTPRD | c.A3117C | p.Q1039H | not detected | not detected | 6.8% | not detected | not detected | yes |
| PTPRT | c.T479C | p.F160S | not detected | not detected | not detected | not detected | 33.8% | no |
| RBM10 | c.A2024G | p.K675R | not detected | not detected | not detected | not detected | 41.7% | no |
| TP53 | c.C9G | p.C3W | not detected | 1.2% | not detected | not detected | not detected | yes |
| TP53 | c.A355G | p.I119V | not detected | 1.7% | not detected | not detected | not detected | yes |
| TP53 | c.G315C | p.M105I | not detected | 1.8% | not detected | not detected | not detected | yes |
| TP53 | c.T316C | p.C106R | not detected | 2.1% | not detected | not detected | not detected | yes |
| ZFHX3 | c.A1604C | p.E535A | not detected | not detected | not detected | not detected | 27.5% | no |
| ZFHX3 | c.A7529G | p.Q2510R | not detected | not detected | not detected | not detected | 39.7% | no |

\*variants were considered pathogenic when more than 50% of the 8 predictors (SIFT, Polyphen2 HVAR, LRT, Mutation Taster, Mutation Assessor, FATHMM, CADD and VEST) considered the alteration as pathogenic/damaging/deleterious/harmful. ND= not defined

**Table 2.**
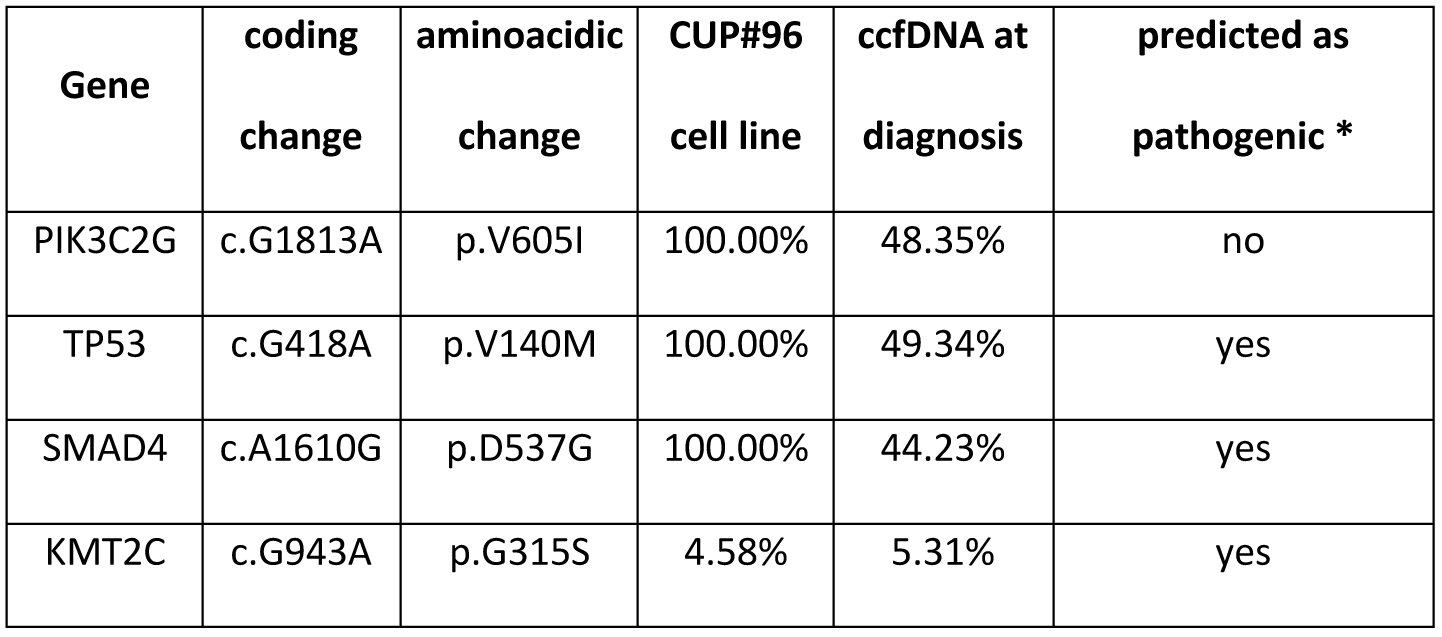
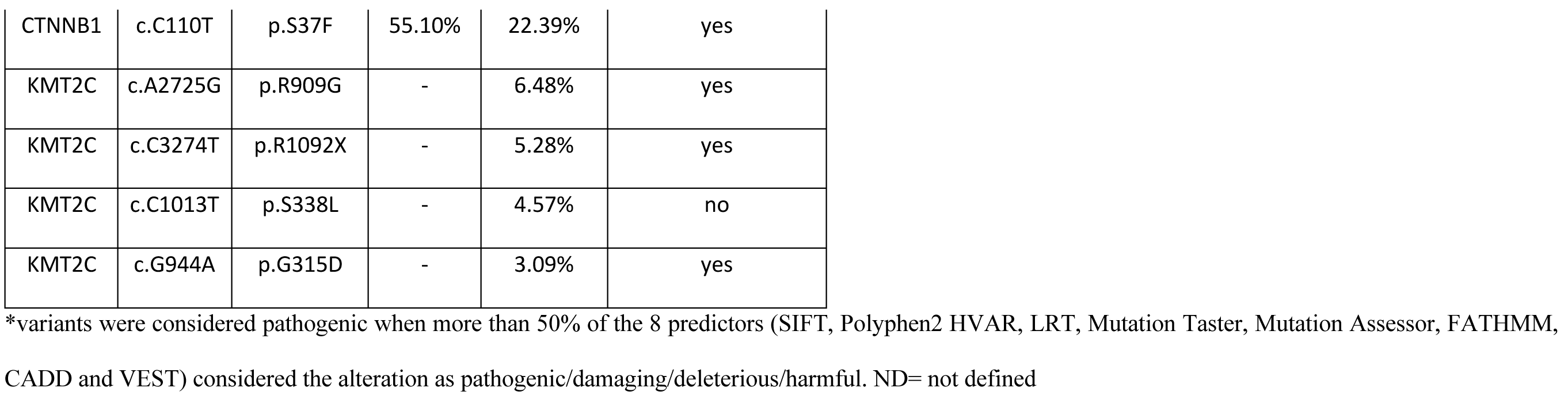
Genetic alterations detected in CUP#96 patient-derived tissues and models.

### Cell type identification using single cell transcriptomics

Single-cell (sc) gene expression profiles of CUP#55 cell line were clustered on the basis of their reciprocal similarity, leading to the identification of two separate clusters (**Figure 3A**). Automated cell type annotation using the scType method identified the liver as the most likely tissue of origin (**Figure 3B**).

**Figure 3.**
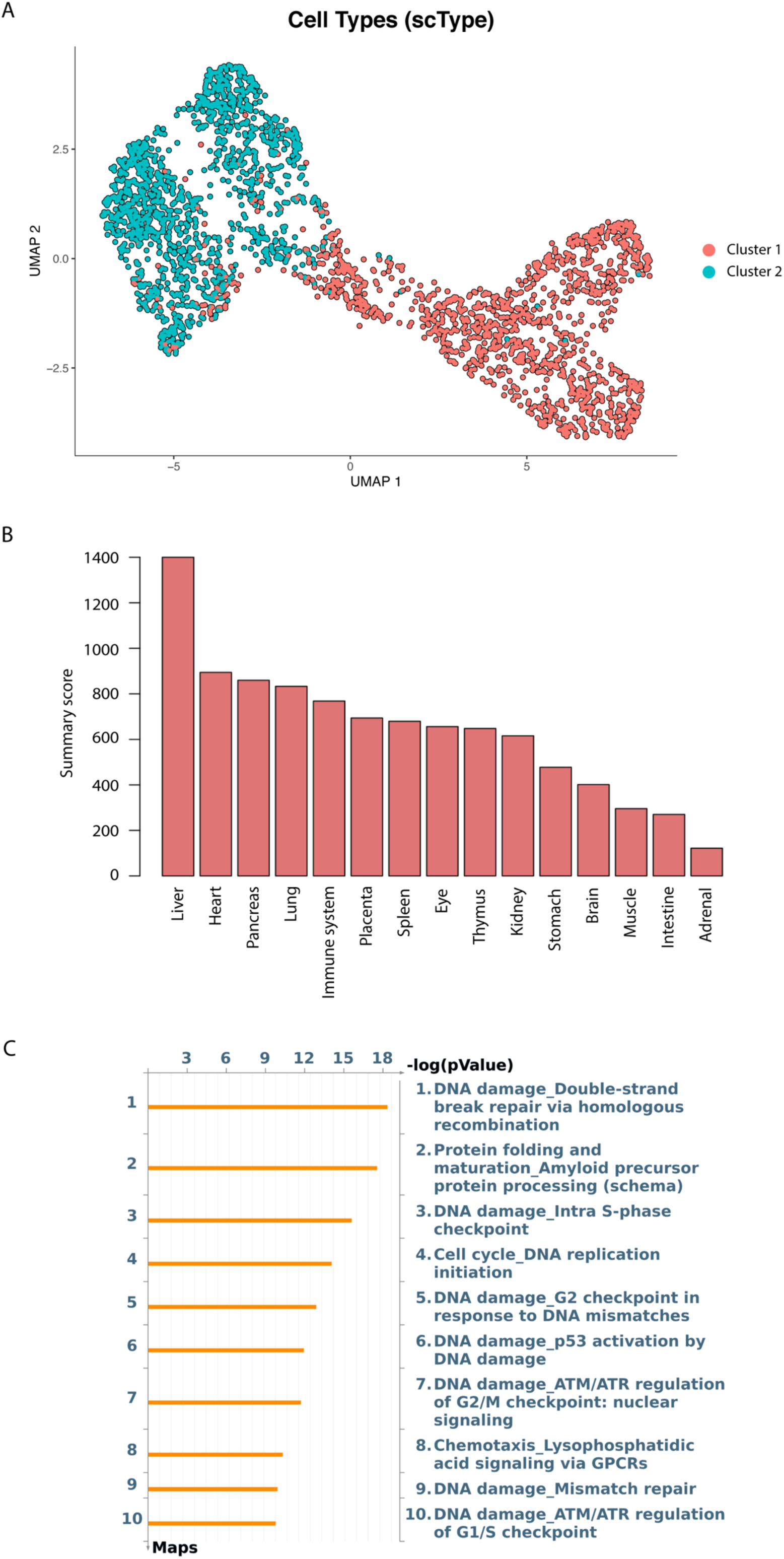
Single cell transcriptome analysis of CUP#55. A) Cell clustering based on the similarity among expression profiles, obtained through the scLCA method; cells are separated in two distinct clusters **B)** Scores computed by the scType method for putative tissues of origin for CUP#55 cells **C)** MetaCore pathway analysis of genes markers of T/NK cells. Differentially expressed genes were imported into MetaCore analytical software to determine the significantly enriched pathways in T/NK cells, which are mostly related to cell division and DNA repair.

We then applied a mapping approach using an atlas of liver cells as a reference, with the objective of inferring the most likely type of origin for CUP cells in each cluster. While 93% of cells in cluster 1 were annotated as cholangiocytes (**Figure S5A**), the classification of cluster 2 was more diverse: specifically, mapped types were split between cholangiocytes (73% of cells) and T/NK cells (27%). As this separation did not correspond to any difference in cell morphology, we double-checked the association manually. We identified the marker genes for the T/NK subgroup (**Supplemental Material file S1**). This list is notable for two reasons: most of the genes are implicated in the cell cycle, and in particular with the mitotic phase and DNA repair, as demonstrated by the results of Metacore pathway enrichment analysis (**Figure 3C**). Furthermore, the list lacks any of the markers which characterize the T/NK group in the liver atlas (CCL5, KLRB1, NKG7, CD69, GZMA, CCL4, CST7, GNLY, GZMK). None of the cells in cluster 2 expresses any of those genes. This suggests that the identification of T/NK cells by reference mapping is likely incorrect and driven by the confounding effect of the cell cycle (**Figure S5B**). Indeed, an average of 46% of the cells in the reference atlas are actively cycling, but when we focus on T/NK cells alone the percentage rises to 74%.

### Expression profiles of cell clusters

The transcriptome analysis highlighted the presence of two separate cell clusters (**Figure 3A**), labelled cluster 1 and cluster 2, with significant differences in their expression profiles. We identified two sets of positive marker genes which characterize each cluster. Specifically, we obtained a total of 1051 markers for the cluster 1 and 3059 for the cluster 2 (**Supplemental Material file S2**). Functional annotation with Metacore of the markers for cluster 1 highlighted an over-representation for pathways including Antigen presentation, Cytoskeleton remodeling and Cell adhesion (**Figure S6**). A similar analysis of cluster 2 markers identified the following enriched pathways: regulation of Wnt/beta-catenin pathway, VEGFR2 signaling and Double-strand break repair.

The sc gene expression analysis confirms the abundant expression of FGFR2 transcript and the variable expression of genes markers of EMT and stemness (**Figure 4**), with SNAI1 and TGFB1 more expressed in cells belonging to cluster 1, and CD44, ZEB1 and HIF1A in cells of cluster 2.

**Figure 4.**
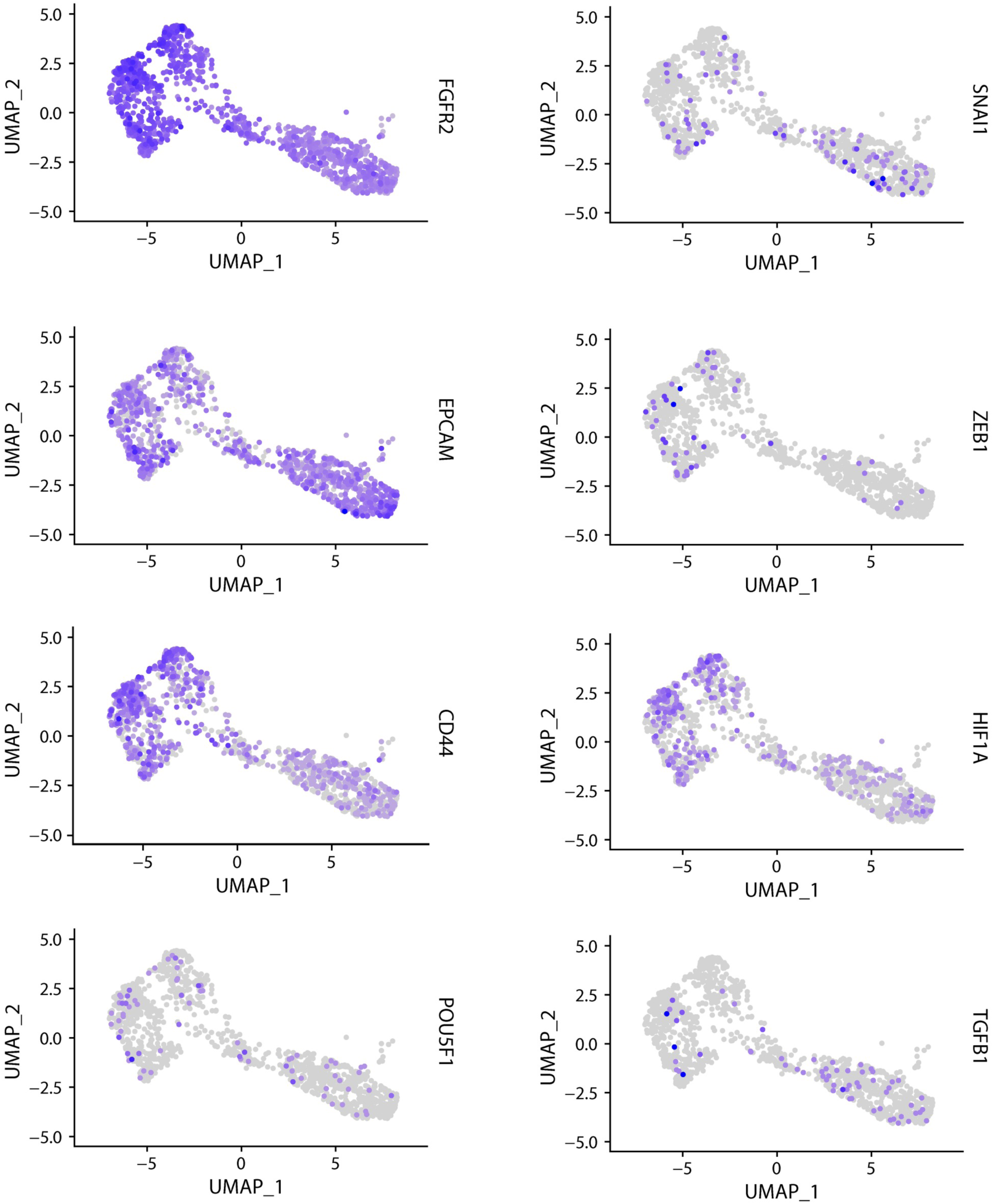
Single cell gene expression. The panels show the expression and distribution in CUP#55 single cells of the FGFR2 gene and other selected markers expressed in liver cancer stem cells and involved in the EMT process.

### FGFR2 amplification in CUP#55 and CUP#96 cell lines provides the rationale to target the receptor using a selective inhibitor

FGFR2 genomic alterations are detected in CUPs with a frequency ranging from 3-4% (**Figure S7**). The *FGFR2* amplification in our two models was confirmed with Fluorescence In Situ Hybridization (FISH) and droplet digital PCR (ddPCR). FISH demonstrated the different nature of *FGFR2* amplification: CUP#96 displays an extrachromosomal DNA amplification in the form of double minutes; CUP#55 shows a homogeneously staining region (HSR) associated with *FGFR2* chromosomal amplification (**Figure 5A**). The copy number variation (CNV) of *FGFR2* gene was confirmed by ddPCR using a probe-based assay in circulating cell-free (ccfDNA), cell line and PDX of both patients (**Figure 5B**). *FGFR2* copy number (CN) in tumor DNA is about 90 for CUP#55, and >400 for CUP#96.

**Figure 5.**
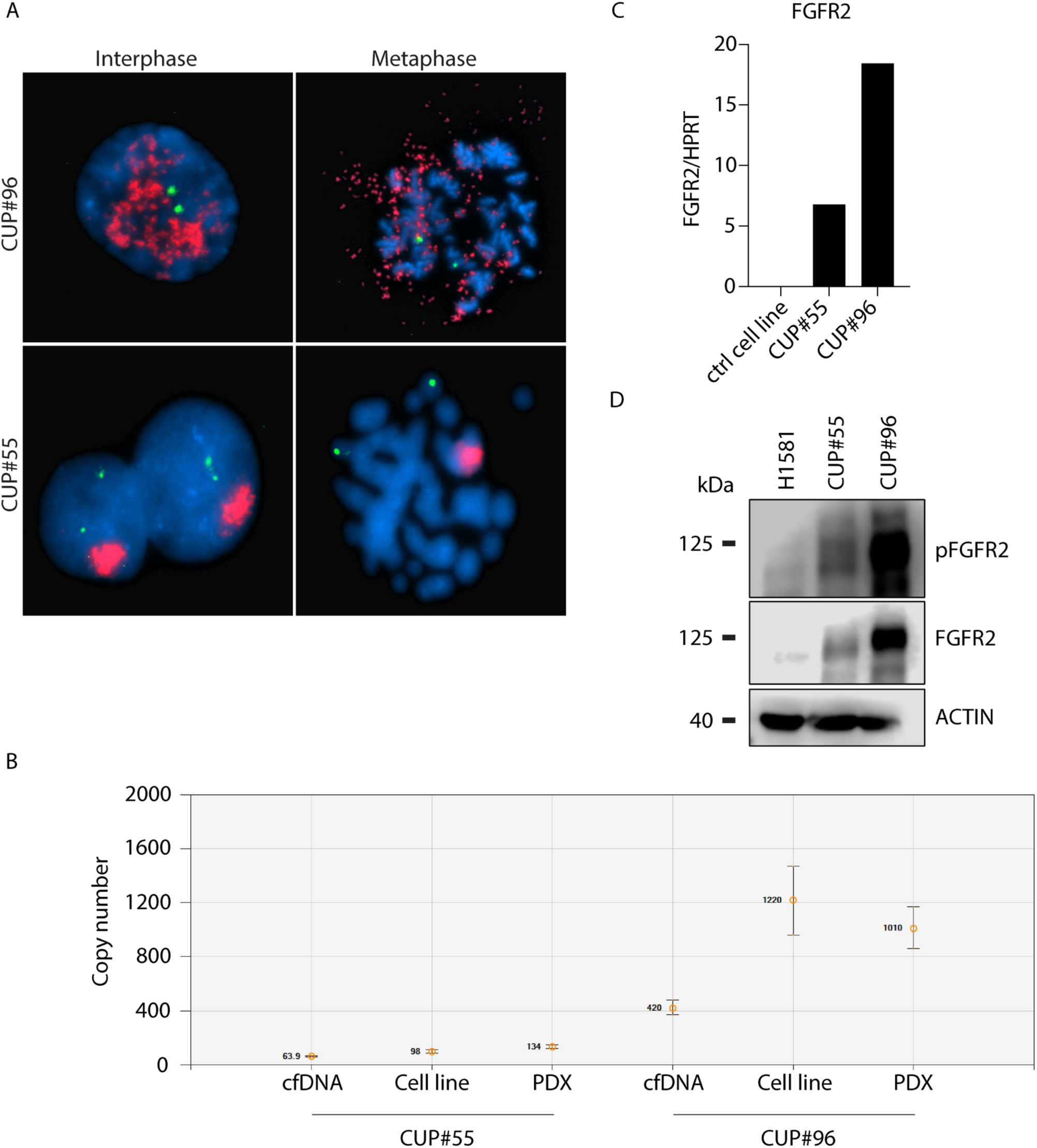
FGFR2 amplification in CUP models. A) FISH analysis shows the different nature of FGFR2 amplification in the two CUP cell models: double minutes for CUP#96 and HSR for CUP#55; FGFR2 gene probe is in red, Chr10 centromere probe in green, nucleus in blue. B) Copy number variation (CNV) analysis of FGFR2 gene detected by ddPCR using a probe-based assay. C) FGFR2 gene expression in the two amplified models and a control not amplified cell line. Bars represent the ratio between the gene copies and the reference HPRT copies assessed by ddPCR. D) Evaluation by western blot analysis of total and phosphorylated forms of FGFR2 in CUP#55 and #96, compared to the H1581 NSCLC cell line.

Consistent with the CN amplification, FGFR2 gene expression is increased in both cell lines (**Figure 5C**). We monitored the FGFR2 isoform expressed by the cell lines and found that both cell lines express the epithelial FGFR2 IIIb isoform and the mesenchymal FGFR2 IIIc isoform (**Figure S8**). Western blot analysis confirmed that CUP#96 cells express higher levels of total FGFR2 protein compared to CUP#55 cells. Interestingly, in both models the FGFR2 appears constitutively phosphorylated (**Figure 5D**). Notably, the FGFR2 phosphorylation/expression is higher than in a reference FGFR2 positive cell line (H1581), derived from a lung adenocarcinoma with FGFR1 and FGFR2 amplification (23) (**Figure 5C-D**). FGFR2 gene amplification and activation in CUP#55 and CUP#96 cells provided the rationale for the pharmacological targeting of this receptor for therapeutic intervention.

### Simultaneous inhibition of FGFR2/AKT and MAPK pathways by BGJ398 and trametinib, respectively, induces synergistic anti-proliferative and pro-apoptotic effects in CUP models *in vitro*

We preliminarily tested a panel of multikinase inhibitors (BJ398, dovitinib, ponatinib) for their effect on CUP#55 cell vitality. We selected BGJ398 (infigratinib), a selective inhibitor of the FGFR family, for its greater antitumor activity (**Figure S9**) for further testing in CUP#55 and CUP#96 cells. In CUP#55, the treatment with this drug at different concentrations (range 0.5-2.5 µM) inhibited FGFR2 phosphorylation and the downstream AKT/mTOR/p70S6K signaling (**Figure 6A**). In contrast, ERK1/2 remained phosphorylated even at the highest BGJ398 concentration. CUP#55 cells were sensitive to BGJ398 treatment, although a complete inhibition of their viability/proliferation was not achieved even at 1 µM BGJ398 concentration, suggesting that FGFR-independent mechanisms sustain the activation of the MAPK pathway and contribute to cell growth in this model. These results prompted us to test whether targeting the MAPK pathway with trametinib, a highly specific inhibitor of MEK1/2 proteins, might improve the efficacy of BGJ398 treatment. The drug combination inhibited both the AKT and MAPK pathways almost completely (**Figure 6B**), thereby evidencing a highly significant synergistic inhibition of cell proliferation, as indicated by the comparison with the theoretical interaction curve in the Bliss experimental model (p<0.001) (**Figure 6C**). Interestingly, even if the single treatment with either BGJ398 or trametinib did not induce cell death in CUP#55 cells, their combination had a cytotoxic effect, as demonstrated by fluorescence microscopy analysis of Hoechst 33342/PI-stained cells; the morphology of the stained nuclei suggested that the cells died by apoptosis (**Figure 6D, Figure S10**).

**Figure 6.**
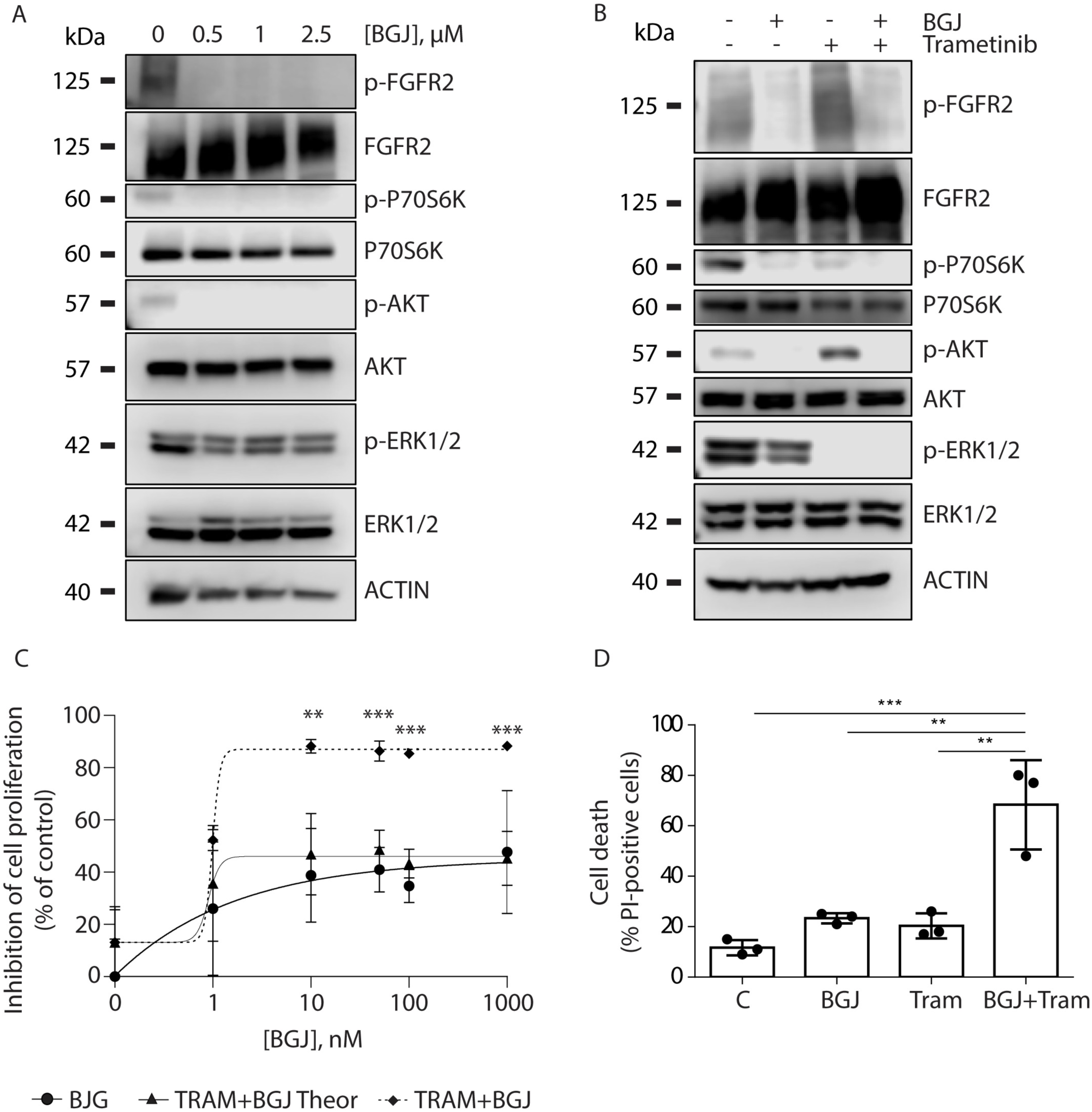
*In vitro* FGFR2 targeting and synergic activity of FGFR2 and MEK inhibitors in CUP#55. A) CUP#55 cells were treated with BGJ398 at the indicated concentrations; after 24h, protein extracts were analyzed by Western blotting for the indicated proteins. The results are representative of two independent experiments; B) CUP#55 cells were incubated with BGJ398 1µM, trametinib 100 nM, or the combination; after 24h, protein extracts were analyzed by Western blotting for the indicated proteins. The results are representative of two independent experiments; C) CUP#55 cells were treated with increasing concentrations of BGJ398 in combination with trametinib 100 nM. After 96h, cell proliferation was assessed by MTS assay. The data are expressed as percent inhibition vs. control. The asterisks indicate the statistical significance vs. the corresponding points of the Bliss Theoretical curve. The results are representative of three independent experiments; D) CUP#55 cells were incubated with BGJ398 1µM and/or trametinib 100 nM; after 96h, the percentage of cell death was evaluated by fluorescence microscopy after Hoechst 33342/PI staining. The data are mean values ±SD of three independent experiments. **p<0.01, ***p<0.001.

Then, we analyzed the effects of FGFR2 inhibition by BGJ398 treatment in CUP#96 cells. Inhibition of FGFR2 phosphorylation by BGJ398 treatment resulted in the downregulation of phosphorylated forms of AKT, p70S6K, and ERK1/2 levels, suggesting that both AKT and MAPK pathways are downstream of FGFR2 in this cell model (**Figure 7A**). We tested the combination of BGJ398 with trametinib also in these cells and found that it produced a remarkable synergistic inhibition of cell proliferation (**Figure 7B-C**), comparable to that observed in CUP#55 cells, suggesting that trametinib can provide a more effective inhibition of the MAPK pathway leading to suppression of cell growth also in this cell model. Of note, BGJ398 and trametinib alone were cytotoxic in CUP#96 cells, an effect that was further enhanced by the combined treatment (**Figure 7D**). Altogether, these findings indicate that trametinib significantly improves the efficacy of FGFR2 targeting in FGFR2-amplified CUP cells.

**Figure 7.**
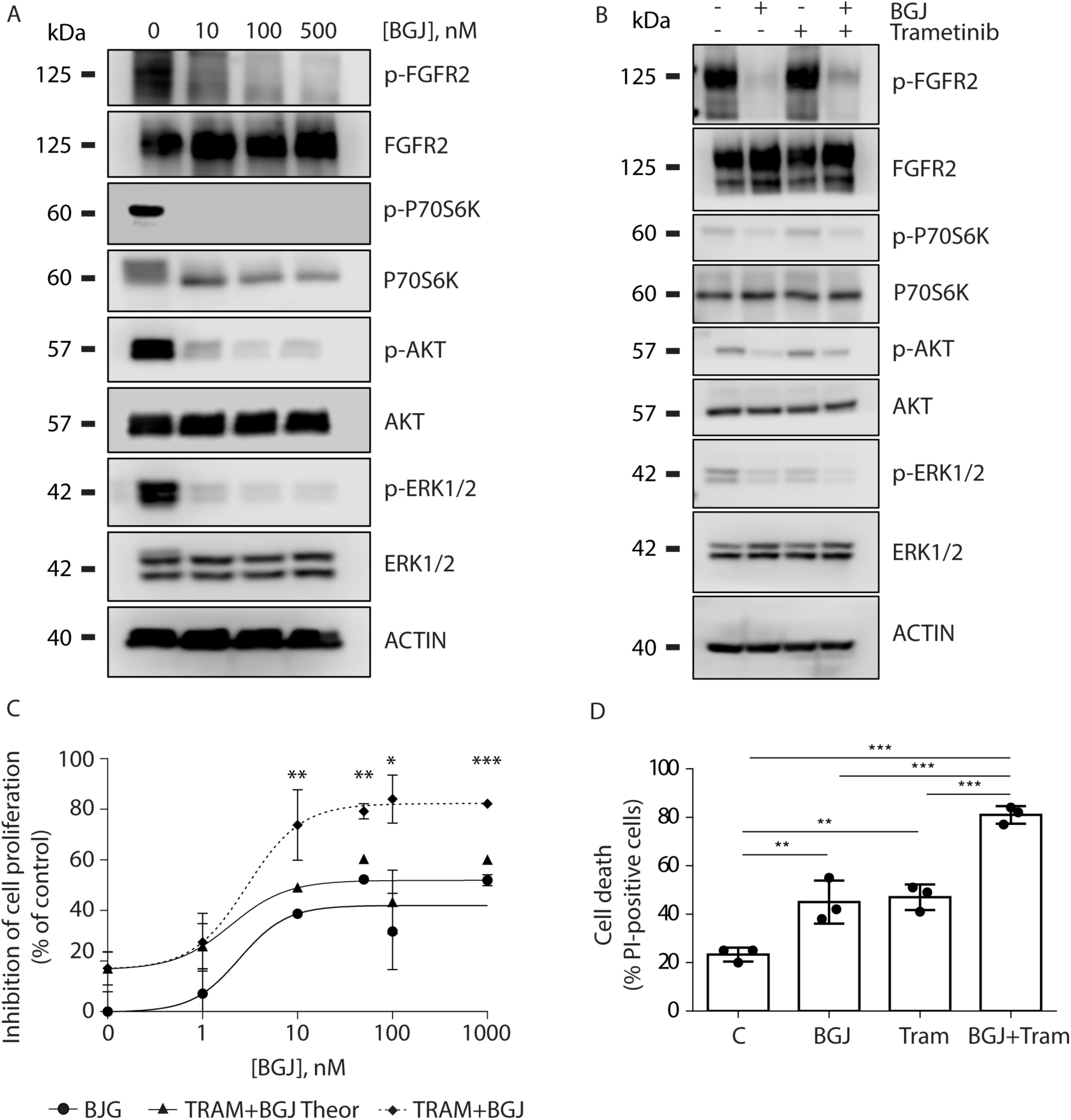
*In vitro* FGFR2 targeting and synergic activity of FGFR2 and MEK inhibitors in CUP#96. A) CUP#96 cells were treated with BGJ398 at the indicated concentrations; after 24h, protein extracts were analyzed by Western blotting for the indicated proteins. The results are representative of two independent experiments; B) CUP#96 cells were incubated with BGJ398 100nM, trametinib 10 nM or the combination; after 24h, protein extracts were analyzed by Western blotting for the indicated proteins. The results are representative of two independent experiments; C) CUP#96 cells were treated with increasing concentrations of BGJ398 in combination with trametinib 10 nM. After 96h, cell proliferation was assessed by MTS assay. The data are expressed as percent inhibition vs. control. The asterisks indicate the statistical significance vs. the corresponding points of the Bliss Theoretical curve. The results are representative of three independent experiments; D) CUP#96 cells were incubated with BGJ398 100 nM and/or trametinib 10 nM; after 96h, the percentage of cell death was evaluated by fluorescence microscopy after Hoechst 33342/PI staining. The data are mean values ±SD of three independent experiments. *p<0.05; **p<0.01, ***p<0.001, ****p < 0.0001.

### *In vivo* synergic activity of FGFR2 and MEK inhibitors

We tested whether in *vitro* findings on FGFR2 and MEK inhibitors synergic activity could be recapitulated in CUP#55 and CUP#96 PDX models *in vivo*.

Mice were treated with control vehicle, BGJ398 (15 mg/kg), trametinib (0.6 mg/kg), and trametinib/BGJ398 combination (N=5 per group). The scheme of different treatments is reported in Figure 8 A and B, and the effect of different drugs was evaluated measuring the tumor volume. Single drug (BGJ398 or trametinib) treatment significantly reduced the tumor volume in PDX#96 but not in PDX#55 xenografts mice if compared to tumors of mice treated with vehicle. However, the combination of trametinib and BGJ398 resulted more effective than the single treatments in both PDX models, as demonstrated by the dramatic reduction of the tumor volume (**Figure 8A, B**). CUP#55 PDX mice suffered from tumor-induced cachexia and for this reason they were sacrificed after 2 weeks of treatment. CUP#96 PDX mice were treated for up to 33 days with the combination and were sacrificed at 6 weeks.

**Figure 8.**
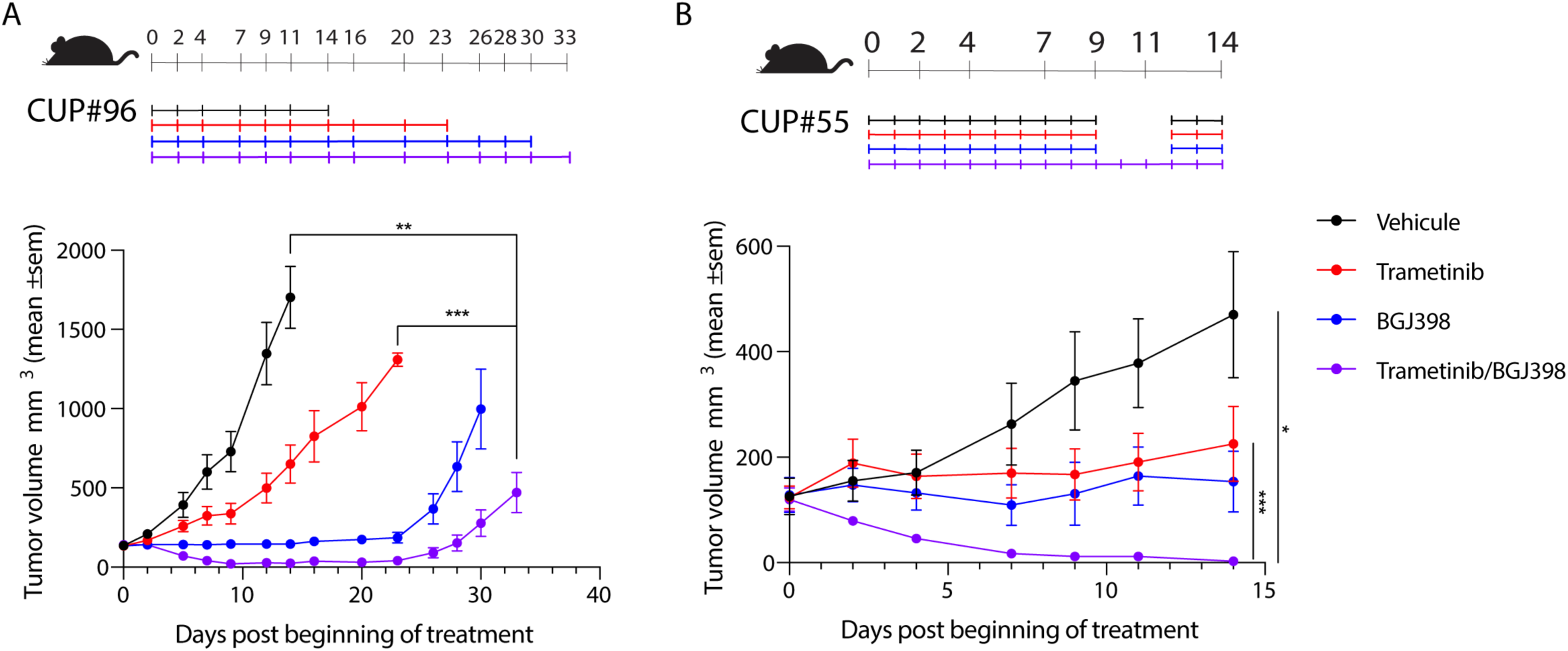
*In vivo* activity of FGFR2 and MEK inhibitors. *In vivo* treatment of A) PDX#96 PDXs and B) PDX#55 PDXs. The timeline shows the day of tumor volume measurement, and the color bars represent the scheme of each treatment. The line graph shows that the combination of trametinib+BGJ398 treatments (purple) is more effective than the single ones (trametinib in red and BGJ398 in blue) in both PDX models. N=5 mice per group. *p<0.05, **p<0.01, ***p<0.001, ****p<0.0001.

Transcriptome shift induced by treatment

We analyzed the early effects of the combined treatment with MEK and FGFR2 inhibitors on CUP#55 transcriptome at the single cell level. The treatment had a strong effect on the transcriptional activity of the two cell clusters: we identified 526 and 2947 differentially expressed genes between the treated and untreated cells, respectively in cluster 1 and 2 (**Supplemental Material file S3**). A gene set enrichment analysis confirmed that the epithelial-mesenchymal transition was influenced by the treatment, although at different degrees in the two clusters. Specifically, the signature identified by Tuan *et al* (24) was the most enriched in cluster 1 (NES -2.36, adjusted p-value 7.81E^-4^), while the signature from Cursons *et al* (25) was the most significant in cluster 2 (NES -3.09, adjusted p- value 3.56E^-8^). The negative sign in both enrichment scores corresponds to an overall downregulation of genes promoting the epithelial phenotype, suggesting a greater impact of the treatment on cells with an epithelial phenotype.

Lastly, we extended our enrichment analysis to all signatures part of msigdb (**Supplemental Material file S4**). We found the signature “ANDERSEN_CHOLANGIOCARCINOMA_CLASS2”, which collects genes overexpressed in cholangiocarcinoma class 2 associated with a poor prognosis, as negatively enriched in cluster 2 (NES -3.34, adjusted p-value 8.53E^-10^). While the enrichment was not significant in cluster 1 (adjusted p-value 0.02), the sign of the normalized enrichment score remained negative (-2.2). Similarly, “HOSHIDA_LIVER_CANCER_SUBCLASS_S1” (genes from ‘subtype S1’ signature of hepatocellular carcinoma, linked to an aberrant activation of the WNT signaling pathway) was enriched with a negative score in cluster 1 (NES -2.4, adjusted p-value 6.1E^-3^), and obtained a negative but not significant score in cluster 2 (NES -2.1, adjusted p-value 0.02). Overall, these results hint at a role of the treatment in reducing the activity of genes involved with cancer progression.

## DISCUSSION

Treatment choice for cancer of unknown primary patients is always challenging due to the inconclusiveness of classical diagnostic investigations in tumor type identification. In this scenario, therapy can be based on empirical approaches, molecular predictions of the primary site, the clinical assessment of the similarity with other known tumor types or more recently, on personalized genetic approaches. Indeed, the combination of molecular and genetic investigations can help the treating clinicians to define the most probable site-of-origin or potential druggable mutations and use this information to choose the best therapeutic strategies.

CUP therapeutic choice has been also hampered by the lack of CUP models on which to test novel therapeutic approaches or investigate the biology of this aggressive disease. Verginelli et al. described the first CUP experimental models adopted to investigate molecular and genetic alterations of the disease (21). The authors demonstrated that CUP models present a specific stem-cell like phenotype and tested for the first time their sensitivity to MEK inhibitors. Here, we described two *in vitro* and *in vivo* models we derived from ascites circulating tumors cells (CTCs) of CUP patients, named CUP#55 and CUP#96. Using a CTC-optimized protocol, we established two long-term cell cultures spontaneously growing as spheroids/tumoroids and expressing stem-cell markers. Combined investigations on microRNA expression, single cell transcriptomics and genomic alterations pointed toward a biliary tract origin for CUP#55 and gastrointestinal origin for CUP#96.

The ascites CTCs were engrafted into mice to generate two patients derived xenografts, which recapitulated the characteristics of the original tumor. The genetic analyses, performed on tumor tissues and models, revealed that the two models shared the FGFR2 amplification as main genetic alteration, although of a different nature. FGFR2 amplification is reported in several solid tumors, including gastric cancer and breast cancer (26), while its translocation is a recurrent feature in cholangiocarcinoma (27).

FGFR2 amplification is associated with increased levels of the protein and its aberrant phosphorylation leads to the activation of downstream pathways, including MAPK-ERK signaling (28), which in turn accelerate cell proliferation. Since FGFR2 amplification is a druggable target (28) we investigated the extent of CUP tumors FGFR2 dependency. The treatment with BGJ398 (infigratinib), a pan-FGFR inhibitor, demonstrated the effectiveness of this target therapy in reducing AKT activation, but did not completely prevent ERK1/2 phosphorylation, which remained active through alternative mechanisms. This finding suggests that FGFR2 targeting in FGFR2-amplified CUPs might not be sufficient to halt tumor growth, due to the concomitant activation of alternative survival/proliferation pathways. Therefore, we investigated the effect of the combined use of trametinib, a MAPK pathway inhibitor selective for MEK1/2. MAPK is a pathway that is frequently activated in CUPs (29), where its activation correlates with worse prognosis. Interestingly, the combined treatment with FGFR2 and MEK inhibitors generated a remarkable synergistic effect, reducing cell growth and viability in both cell models with induction of cell death.

The short-term combined treatment of cultured cells revealed a degree of plasticity in the apparently homogeneous cell line, which could be evidenced only from single-cell transcriptomic data. The transformed cholangiocytes of CUP#55 cell line were differentially affected by the combined treatment, with genes involved in cancer progression being silenced in both cell subgroups (clusters) and cells expressing an epithelial phenotype being more susceptible to treatment-induced cell death. To validate the results in a preclinical model, the corresponding PDXs were treated with the drug combination and the effect was a drastic reduction of the tumor size if compared with single treatments. The treatment combination was well tolerated by the CUP#96 model but aggravated the mice frailty in CUP#55 model, which already presented signed of tumor-induced cachexia.

In the last years, pemigatinib and infigratinib, both orally active agents targeting FGFR1-4, have received an accelerated approval by US Food and Drug Administration (FDA) for the treatment of adult patients with previously treated and unresectable or metastatic cholangiocarcinoma harboring FGFR2 fusion or other rearrangement. Based on some concerns, European agency (EMA) granted approval with the same indication only for pemigatinib. Likewise, erdafitinib, an orally active small potent TKI of FGFR1–4, was granted accelerated approval for patients with locally advanced or metastatic urothelial carcinoma, with susceptible FGFR3 or FGFR2 genetic alterations.

FGFR inhibitors have also been largely tested in gastric cancer patients with less enthusiastic results so far. However, positive results from a phase II trial have been reported by using bemarituzumab in addition to chemotherapy as first line treatment for metastatic HER-2 negative gastroesophageal cancer patients with FGFR2b hyperexpression or FGFR2 gene amplification (30). As such, several phase II and phase III trials currently ongoing. Other new compounds, alone or in combination with other agents, are under investigation for the treatment of multiple solid tumors carrying FGFR alterations. Our results suggest that the combination of FGFR and MEK inhibitors could be a potential strategy to improve clinical outcomes in cancer patients carrying FGFR gene alterations. This means that co-targeting cross-talking pathways may potentiate FGFR inhibition, and improve the therapeutic benefit, as we have demonstrated with the MEK inhibitor trametinib.

Based on the evidence that MEK inhibition in KRAS-mutant lung cancer leads to compensatory MAPK pathway reactivation through FGFR1, combining trametinib with FGFR1-specific inhibitors encapsulated in nanoparticles allowed to efficaciously inhibit growth and proliferation in KRAS- mutant/FGFR compensatory cancer cells (31). In this regard, a phase 1/2 study is recruiting patients with advanced cancer of any tumor type (Part 1) or non-small cell lung cancer (NSCLC) with a confirmed KRAS mutation (Part 2) aiming at determining the recommended dose and antitumor activity of futibatinib (a selective, irreversible FGFR1-4 inhibitor) in combination with binimetinib, a known MEK inhibitor (NCT04965818).

Our study contributes to fill the gap in CUP model availability, through the generation of two human cell lines and corresponding PDXs. We demonstrated how the molecular characterization and genetic profile of the tumor can provide information to predict the most probable site-of-origin and identify druggable targets. In fact, the development of models that mirror the phenotype and genotype of human tumors *in vitro* and *in vivo*, has become a helpful tool for drug screening, particularly to assess new therapeutic combinations that could be translated to patients with the same genetic alterations or molecular features. Finally, the availability of CUP models for future studies will contribute to deepen our knowledge on the mechanisms at the base of CUP high proliferative and metastatic potential and still mysterious biology.

## MATERIALS AND METHODS

### Study approval

Two patients with a confirmed diagnosis of cancer of unknown primary based on ESMO criteria (CUP#55 and CUP#96) were enrolled in this study at the Oncology Unit of IRCCS Azienda Ospedaliero-Universitaria di Bologna (Bologna, Italy). The study was approved by the local ethical committee (Comitato Etico Indipendente dell’Azienda Ospedaliero-Universitaria di Bologna, Policlinico S.Orsola Malpighi) with protocol number EM435-2022_130/2016/U/Tess/AOUBo. All subjects provided a written informed consent to the study participation, in accordance with the Declaration of Helsinki.

### Cell culture

NSCLC cell lines H1581, A549, H460, H2228, H1299 and SK-MEL-28 were from ATCC (Manassas, VA, USA).

Cells were cultured in RPMI-1640 medium (Corning) supplemented with 10% fetal bovine serum (Corning) and maintained under standard cell culture conditions at 37°C in a water-saturated atmosphere of 5% CO2 in air.

### Patients and ascites CTC isolation

About 100 ml of ascites from both patients were collected and immediately processed. Ascites samples were centrifuged at 600 g for 10 minutes at 4°C and the obtained pellet was treated with 10 ml red blood cell lysis buffer (Miltenyi Biotec) for 15 minutes at room temperature. Then, 10 ml of specific culture medium (DMEM/F12 50:50, 2 mM glutamine, 5% Horse serum, hydrocortisone 1 ug/ml, insuline 10 µg/ml, 1X PEN/STREP, EGF 10 ng/ml) was added and centrifuged at 600 g for 10 minutes. The cells were plated in T25 flask with 5 ml of medium in humidified 37 °C/5% CO2 incubator. Medium was changed every 3–4 days. After 10 days, tumor cells begun to grow as tumoroids (CUP#96) and spheroids (CUP#55).

Organoids and aggregates were propagated in 24 well-plates to monitor growth and every 3-4 days splitted 1:2. Long-term CTC cell lines were established: i.e. the cell lines were still viable after thawing cycle and showed a stable growth in medium without supplement (32).

### Pathological and immunofluorescence analysis

Cell lines, human and PDX tumor tissues were fixed in 4% paraformaldehyde for 20 minutes, embedded in paraffin and frozen at -20°C for optimal cutting. Sections of 4 μm were mounted on positively charged microscope slides. Sections were deparaffinized in xylene and rehydrated in graded alcohol. Antigen enhancement was done by incubating the sections in Concentrated Antigen Retrival Solution Citra Plus (BioGenex #HK080-5K) (1:10) as recommended by the manufacturer and blocked with Ventana Antibody Diluent with Casein (Roche-Ventana) for 30 minutes. For immunohistochemistry: Rabbit Monoclonal Primary Antibody Cytokeratin 7 (SP52) (Roche-Ventana) and Rabbit Monoclonal Primary Antibody Cytokeratin 20 (SP33) (Roche-Ventana), were used at 1:2 dilution for cell lines, human and PDX tumor tissues staining.

For immunofluorescence experiment: Mouse Monoclonal Primary Antibody CD44 (or HCAM) sc-7297 (SantaCruz) 1:100 dilution and Mouse Monoclonal Primary Antibody EpCAM FITC (REA764) (Miltenyi) 1:100 dilution was used for cell lines staining. All the antibodies were incubated overnight at 4°C. AbCAM Goat Anti-Rabbit IgG H&L (DyLight® 594) (ab96885) and Donkey Anti-Mouse IgG H&L (DyLight® 488) (ab96875) labeled secondary antibodies were used at 1:100 dilution and incubated 1h. Nuclei were counterstained with 1 μg/mL of Hoechst 33342 (Life Technologies). Confocal images were acquired on Nikon A1 confocal laser scanning microscope, equipped with a 60X, 1.4 NA objective and with 405, 488 nm laser lines.

### Genetic analysis

We used a custom 1.2-Mb SureSelect capture bait library (Agilent Technologies, Santa Clara, CA, United States) for the target enrichment of 92 genes (panel description in (Laprovitera et al., 2021c)). Briefly, libraries were prepared using 50 ng of gDNA input following SureSelectXT HS/SureSelectXT Low Input Target Enrichment with Pre-Capture Pooling protocol (G9702-90005, v. A0, June 2019, Agilent Technologies) and sequenced on NextSeq 500 (Illumina) platform using High Output 2 × 75-bp flow cells. Variant calling and paired analyses (tumor vs. normal) were performed using SureCall software (v. 4.2), applying a filter for tumor/normal tissue/models at 5% and for ccfDNA at 1%. Variants were annotated using ANNOVAR (Wang et al., 2010) and filtered to keep somatic exonic non-synonymous single-nucleotide variants (SNVs), insertions, deletions, multiple nucleotide variants, or long deletions not detected in the normal sample that presented an allele frequency in Non-Finnish European (NFE) population lower than 0.5% (Genome Aggregation Database, GnomAD; Karczewski et al., 2020) and a coverage higher than 100. Bioinformatic pathogenicity prediction, reported in Tables 1 and 2 of the identified variants was performed consulting the prediction score/outcome of 8 prediction models: SIFT (Sort Intolerated From Tolerated; Vaser et al., 2016), Polyphen2 HVAR (Polymorphism Phenotyping v2; Adzhubei et al., 2010), LRT (Likelihood Ratio Test; Chun and Fay, 2009), Mutation Taster (Schwarz et al., 2014), Mutation Assessor (Reva et al., 2011), FATHMM (Functional Analysis Through Hidden Markov Model; Shihab et al., 2013), CADD (Combined Annotation Dependent Depletion; Kircher et al., 2014), and VEST (Variant Effect Scoring Tool; Douville et al., 2016). Variants were considered pathogenic when more than 50% of the above-mentioned predictors indicated it as pathogenic/damaging/deleterious/harmful.

### Single cell transcriptome analysis

The transcriptome of CUP#55 cell line was characterized at the single-cell level. To this end, we relied on the 10X Single Cell 5’ R2-only kit. The resulting libraries were sequenced with the Novaseq 6000 (Illumina) platform using a 2 × 100-bp flow cell, obtaining a total yield of 240 million reads. Gene expression profiles of different cells were quantified by the CellRanger pipeline (version 7.1.0), using as a reference the GENCODE annotation (version 40). The software provided an initial estimate of 23,994 sampled cells. Gene counts were filtered by means of the CellBender method (33) to remove the confounding effects of ambient RNA molecules and random barcode swapping. Around 7% of all droplets were identified by scDblFinder (34) (version 1.14.0) as potential doublet artifacts and were excluded from the rest of the analysis. We also discarded cells characterized by a high proportion of reads from the mitochondrial genome, a signal usually associated with cell membrane rupture and apoptosis. We set our filtering threshold to 50% following the indications of other studies (35) that have observed very high mitochondrial content in hepatocytes. As a last QC filter, we dropped all cells showing a detectable level of expression for less than 500 genes. The final working set consisted of 1437 cells.

Gene expression was loaded in the R environment using Seurat (36) (version 4.3.0.1) and normalized with the sctransform (37) method (v2) to account for gene overdispersion. Cells were clustered using the Latent Cellular Analysis method (38) (scLCA package, version 0.0.0.9000), which combines a cosine**-**similarity measure with a graph**-**based algorithm to automatically identify distinct cell subpopulations. Automatic inference of the tissue type was performed with scType (https://doi.org/10.1038/s41467-022-28803-w) (version 1.0). Reference-based classification of cells was then performed using the Azimuth software (36) (version 0.4.6) and relied on its integrated atlas of the human liver..

We applied the MAST software (39) to identify marker genes of cell clusters. This method (version 1.24.0) implements a generalized linear model specifically designed to account for nuisance variation in scRNA-seq data. We filtered differential expression calls by setting an FDR threshold of 10^-3^, and we also discarded all genes whose log2 fold-change lied below 0.2.

Functional analysis of the resulting gene lists was performed using MetaCore analytical software (Clarivate).

### Gene expression analysis

RNA was extracted from 2 CUP cell lines and 5 cancer cell lines (A549, H2228, H460, H1299, SKMEL). 500ng of RNA for each cell line were reverse-transcribed to cDNA using iSRCIPT cDNA Synthesis Kit (Cat. No. 1708891 Bio-Rad, USA). The expression of CD44, NANOG, OCT4, and FGFR2 was quantified using the QX200 Droplet Digital PCR system (Bio-Rad, Hercules, CA, USA). Bio-Rad EvaGreen protocol for gene expression quantification was used to quantify the gene copies per ng of cDNA by QuantaSoft Analysis software (Bio-Rad, Hercules, CA, USA). Gene/HPRT (reference gene) ratio was used to normalize the gene expression. The primers used for the gene expression analysis are reported in supplementary material.

### Fluorescence In-Situ Hybridization

**(FISH)** FISH analysis was performed on fixed CUP#55 and CUP#96 interphase nuclei and metaphases. Two Empire Genomics probes (Empire Genomics, Buffalo, NY) were employed following the manufacturer’s protocol. Specifically, the FGFR2 gene probe (orange) maps on chr:10q26.13 and the CEP10 probe maps to the centromeric region of Chromosome 10. CUP#55 and CUP#96 cells were counterstained with 40,6-diamidino-2-phenylindole (DAPI) for nuclear detection. Analysis was performed using Olympus BX53 microscopy equipped with the appropriate filter sets and CytoVision software (Leica Biosystems, Nussloch, Germany).

### Detection of *FGFR2* amplification

Total DNA was extracted from CUP#55 and CUP#96 cell lines and PDXs using QIAamp DNA Mini Kit (Cat. No. 5130450, Qiagen). Cell free DNA was extracted from 1mL plasma with Maxwell RSC ccfDNA Plasma Kit (Cat No: AS1480, Promega). Tumor DNA was extracted using the QIAamp DNA FFPE Tissue Kit (Cat No: 56404, Qiagen, Hilden, Germany). To quantify the copy number of FGFR2 in all samples, a probe-based droplet digital PCR assay was used. RPP30 was tested as reference gene for diploid copy number. RPP30 probe (dHsaCP2500350, Bio-Rad) was labeled with HEX, and FGFR2 probe (dHsaCB2500320) was labeled with FAM. Droplet digital PCR was performed with the QX200 Droplet Digital PCR system (Bio-Rad, USA) as described in (10). FGFR2 gene copy number was calculated by QuantaSoft Analysis software (Bio-Rad, Hercules, CA, USA) as FGFR2/RPP30 ratio.

### Cell Viability and Drug response assays

Cell viability/proliferation was evaluated by a CellTiter-Glo 96 Aqueous One solution Assay (Promega). Cell death was analyzed by fluorescence microscopy after staining with Hoechst 3342 and Propidium Iodide (PI) (40). BGJ398 (infigratinib) and trametinib (mekinist) were purchased from Selleckchem (Houston, TX), and dissolved in DMSO. DMSO concentration never exceeded 0.1% (v/v); equal amounts of the solvent were added to control cells. The effect of the drug combination was evaluated using the Bliss interaction model (41).

### Single cell transcriptome of treated cells

We profiled the transcriptome of CUP#55 cells treated with MEK and FGFR2 inhibitors for 12 hours using a single-cell protocol similar to the one discussed for the CUP#55 line. Illumina sequencing resulted in 260 million reads, providing a reliable readout for 1789 cells after the application of quality control filters. The shift in gene expression between treated and untreated cells was evaluated separately for each cluster using MAST and setting an FDR cutoff of 10^-6^.

We employed fgsea (version 1.24.0) (42) to perform a gene set enrichment analysis. Gene signatures were retrieved using msigdb (Bhuva D, Smyth G, Garnham A (2022). _msigdb: An ExperimentHub Package for the Molecular Signatures Database (MSigDB) 10.18129/B9.bioc.msigdb package version 1.6.0, database version 7.5). We downloaded an additional collection of gene sets specifically related to the epithelial-mesenchymal transition from the EMTome database (43) (data retrieved on 05/03/2023). We finally filtered fgsea results by placing an upper threshold of 10^-2^ on the Benjamini-Hochberg adjusted p-values.

### Western blotting

For Western Blot analysis, 8×10^5^ cells from CUP#55 or CUP#96 were seeded in 6 well-plates in complete culture medium. At the end of the treatments, the cells were harvested and centrifuged. The procedures for protein extraction, solubilization, and protein analysis by western blotting are described elsewhere (44). Antibodies against p-FGFR (#3471), FGFR2 (#11835), p- ERK1/2^Thr202/Tyr204^ (#4370), ERK1/2 (#4695), p-AKT^Ser473^ (#9271), AKT (#9272), p-P70S6K^Thr389^ (#9205), P70S6K (#9202) were from Cell Signaling Technology, Incorporated (Danvers, MA); anti-β-actin (clone B11V08) was from BioVision (Milpitas, CA). Horseradish peroxidase-conjugated secondary antibodies and the chemiluminescence system were from Millipore (Millipore, MA). Reagents for electrophoresis and blotting analysis were from BIO-RAD Laboratories (Hercules, CA). The chemiluminescent signal was acquired by C-DiGit R Blot Scanner and the bands were quantified by Image StudioTMSoftware, LI-COR Biotechnology (Lincoln, NE).

### Growth of CUP tumors in immunocompromised mice

PDX studies were run at XenTech in compliance with authorization n. APAFIS#30365- 2021012215599431 v1 conferred from the French Ministry of Agriculture and Food. The authorization to use animals in the CERFE facility (Evry-Courcouronne, France) was obtained by The Direction Départementale de la Protection des Populations, Ministère de l’Agriculture et de l’Alimentation, France “Direction of the Veterinarian Services, Ministry of Agriculture and Food, France” (agreement No. D-91-228-107).

An approximate number of 3-4000 cells or 30-40 organoids were resuspended in 100 µl of culture medium, diluted 1:1 in matrigel and grafted in the interscapular region of NOD/Scid-IL2Rγ-/- (NSG) or NOD/Shi-scid/IL-2Rγnull (NOG) mice. Tumor fragments were sampled from the resected tumor, minced on ice and immediately placed in MACS Tissue Storage Solution (Miltenyi Biotech) and transferred to the animal facility, fragmented and grafted in the interscapular region of Athymic Nude-Foxn1nu mice. Mice were monitored twice weekly for signs of tumor growth. Tumor growth from first implantation occurred in 135 days for CUP#55 and 40 days for CUP#96. Growing tumors were serially transplanted onto recipient mice and the fragments of the tumor harvested for IHC analysis, DNA and RNA extraction. To immortalize each PDX, vials of tumor fragments at different passages were placed in 90% FCS/10% DMSO or glycerol, and stored at -150°C.

### *In vivo* treatments

Treatment efficacy study on each PDX were run as follows. Tumor fragments of the same passage were transplanted subcutaneously onto 3-24 mice (donor mice). When the tumors reached 700 to 1764 mm^3^, donor mice were sacrificed, and tumors were cut into fragments measuring approximately 20 mm^3^. Mice aged 8 to 11 weeks were anaesthetized with 100 mg/kg ketamine hydrochloride and 10 mg/kg xylazine, and one tumor fragment was placed in the interscapular fat pad. On the day of enrollment in the study mice with tumor volume ranging 60 to 200 mm^3^ were randomly attributed to the different groups:

From day 12 to day 14 half dose was administered due to body weight loss.

Trametinib was purchased at Carbosynth and suspended in 10% DMSO; 40% PEG300; 5% Tween-80; 45% NaCl 0.9% for administration; BGJ398 (infigratinib) was purchased at MedChem Express and diluted in 50% Acetic acid/acetate buffer / 50% PEG300 for administration. Tumor volume was evaluated by measuring tumor diameters with a caliper, three times a week during the treatment period (from D0 to D27); all animals were weighted, and tumor size measured the same day. During the whole experimental period, animals were monitored every day for physical appearance, behavior, and clinical changes.

## Data availability statement

Processed single-cell transcriptomic data are available as supplementary information. MicroRNA, genomic and single-cell transcriptomic raw data are available from the authors upon request.

## Acknowledgments

The research leading to these results has received funding from Fondazione Italiana per la Ricerca sul Cancro AIRC grant IG 2021 - ID. 25789 to Manuela Ferracin, Pallotti Institutional funds to Manuela Ferracin, Fondazione del Monte to Manuela Ferracin, UNIPD PRID 2019 – ID. BIRD193023 to Gabriele Sales.

## Author contributions

Conceptualization: AC, MF Methodology: EP, SC, GS, AC

Investigation: AC, CF, IS, NL, GG, MR, MN, SS, EB, GD, RR

Funding Acquisition: MF

Resources: FG, KR, ML, IG, AD, OD, PGP, AA, GS and MF

Writing-Original Draft: AC, IS, CF, GS, MF Writing-Review & Editing: GAC, MB, AA, GS, MF

## Supplemental Figures titles and legends

**Figure S1.**
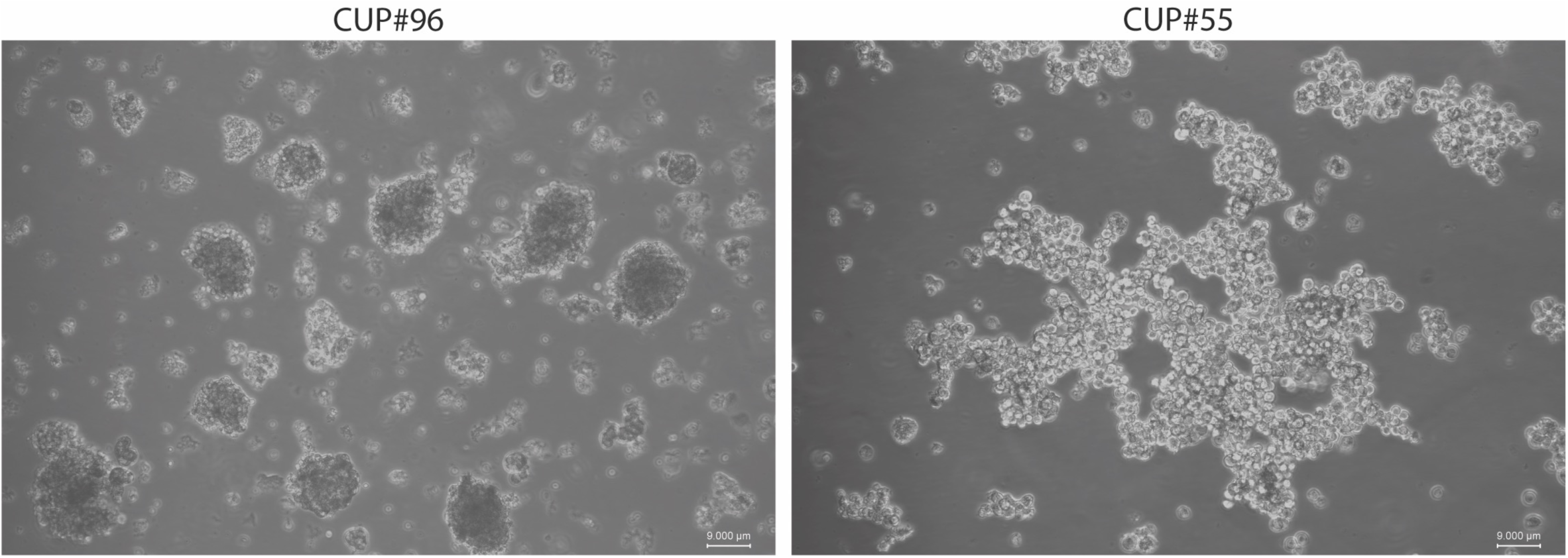
Brightfield representative images of cultured cell lines growing as in suspension 3D- structures. CUP cell lines display discrete *in vitro* growth characteristics. CUP#96 grow as tightly compact spheroids and require trypsinization for passaging. CUP#55 forms loosely aggregated and weakly tight strands requiring only gentle mechanical dissociation for disaggregation. All established cell lines remain viable after freezing and thawing at various passages (self-renew more than 20 passages). Scalebar: 9000um.

**Figure S2.**
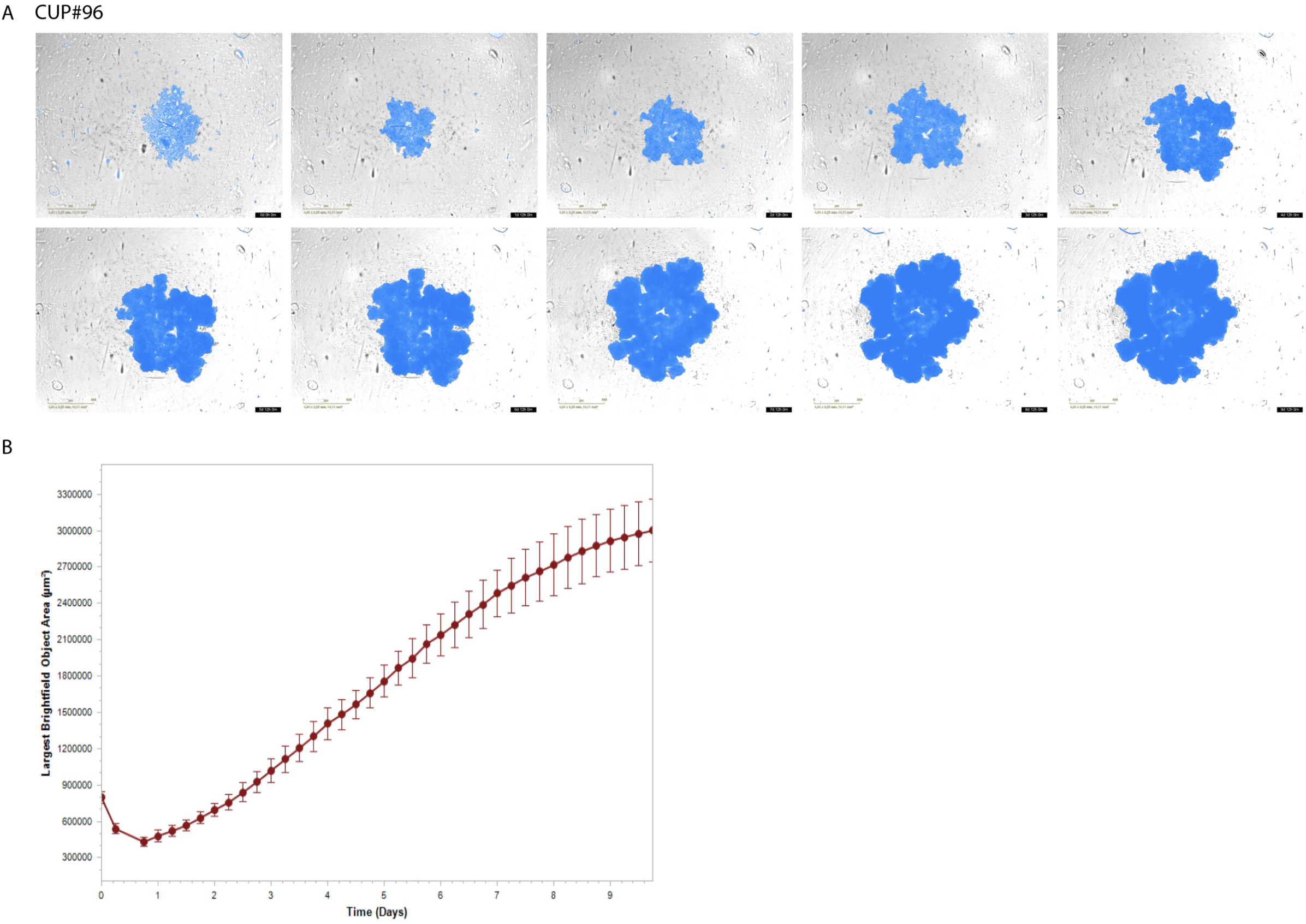
CUP#96 cell growth. A) Panel of brightfield images acquired every 24h for 10 days, in blue the mask used for the object area measurements. B) Growth curve obtained for CUP#96 spheroid analyzing images acquired every 6 hours.

**Figure S3.**
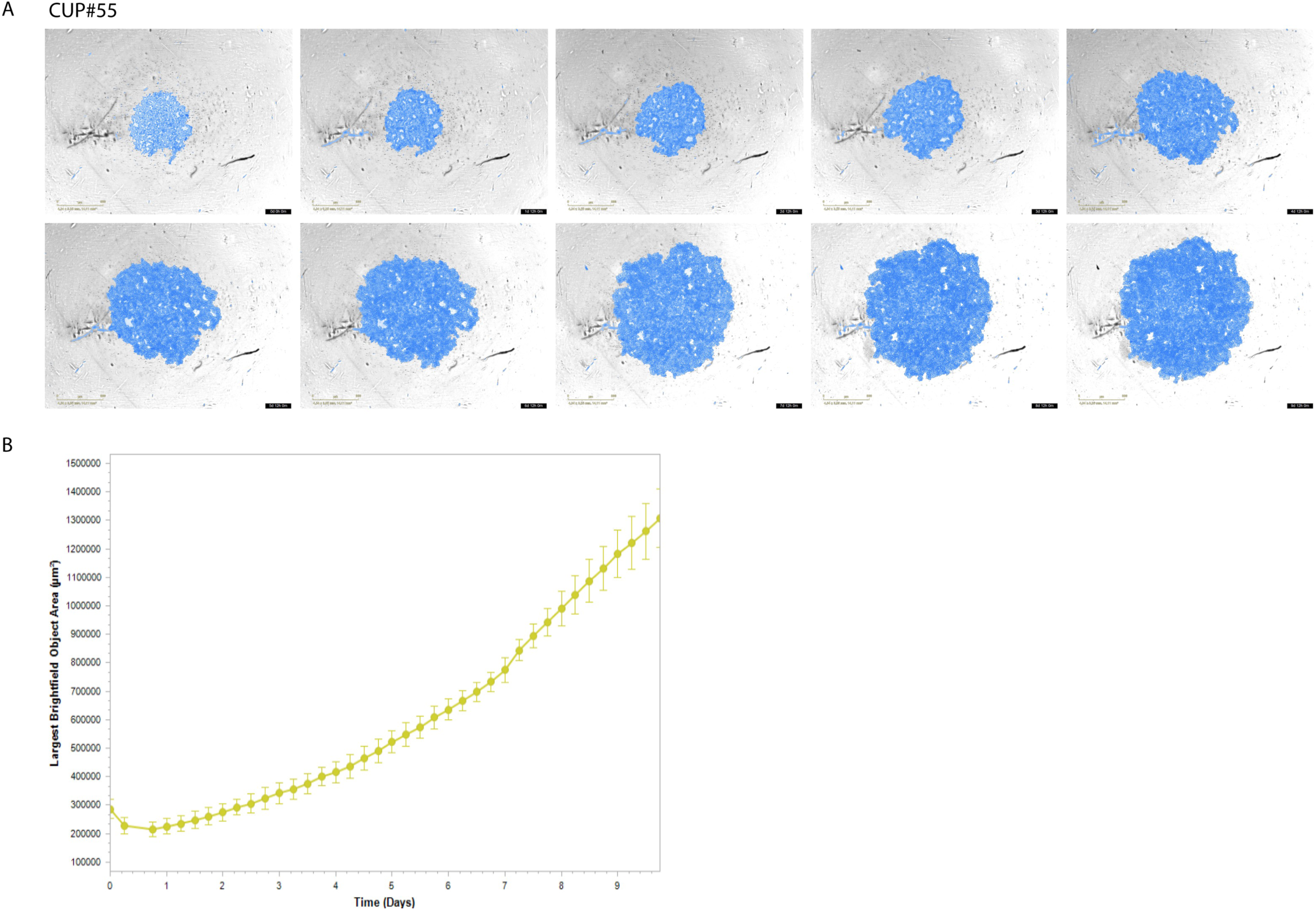
CUP#55 cell growth. A) Panel of brightfield images acquired every 24h for 10 days, in blue the mask used for the object area measurements. B) Growth curve obtained for CUP#55 cluster analyzing images acquired every 6 hours.

**Figure S4.**
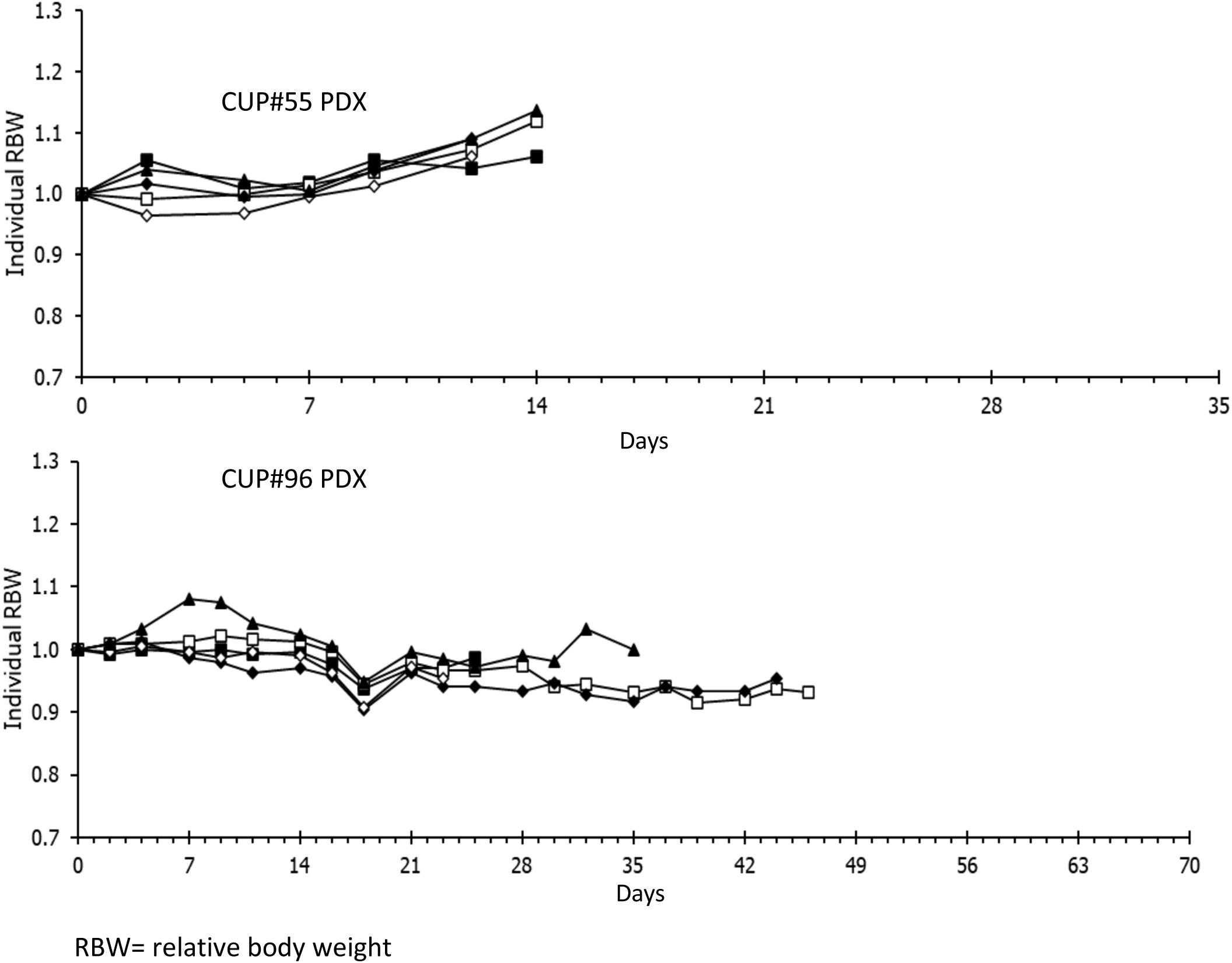
Relative body weight (RBW) graph. Graph showing the RBW over time of N=5 mice for CUP#96 (upper panel) and CUP#55 (lower panel) PDX models. CUP#96 PDX showed a regular RBW for the entire duration of treatments instead CUP#55 PDX were sacrificed after 14 days due to cachexia induced by the tumor.

**Figure S5.**
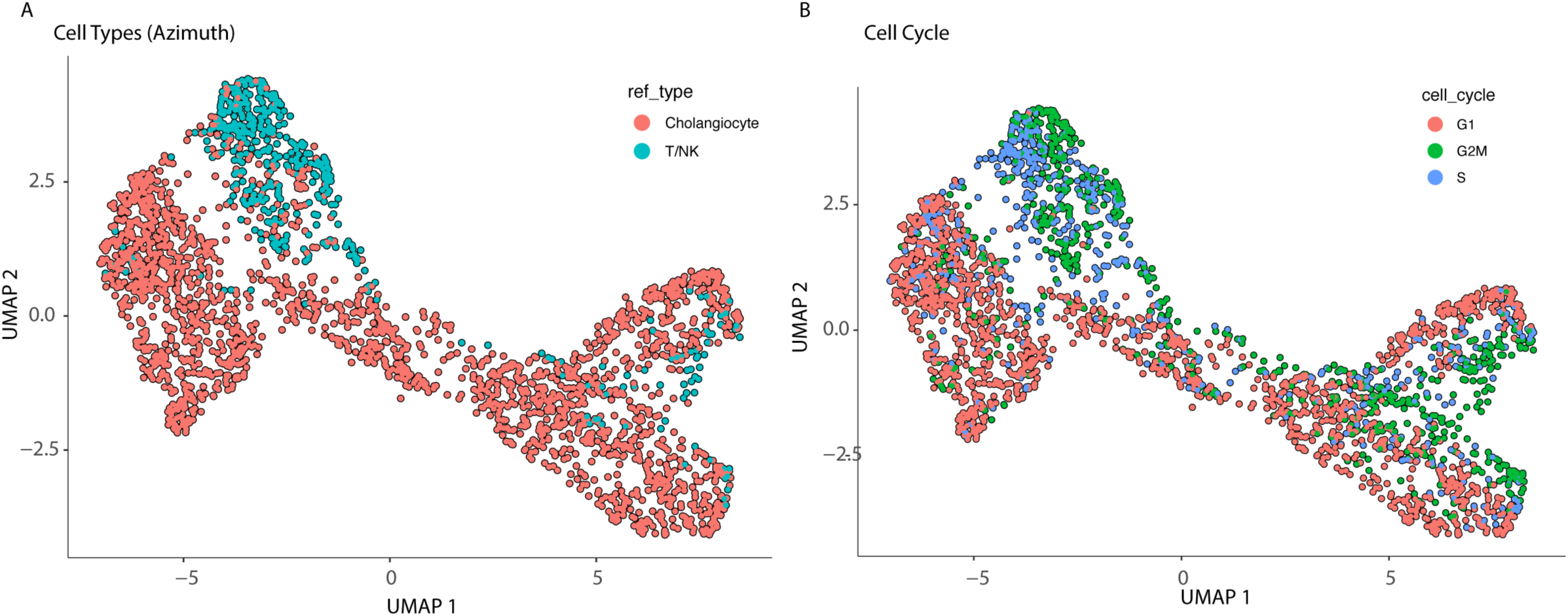
FigureS5 Single cell transcriptomic analysis of CUP#55. A) Cell type identification applying a reference-based classification based on a liver cell atlas (Azimuth software) showed two distinct subgroups: cholangiocytes and T/NK cells B) Cell cycle phase of single cells: cell cycle has a cofounding effect in the identification of the T/NK subgroup.

**Figure S6.**
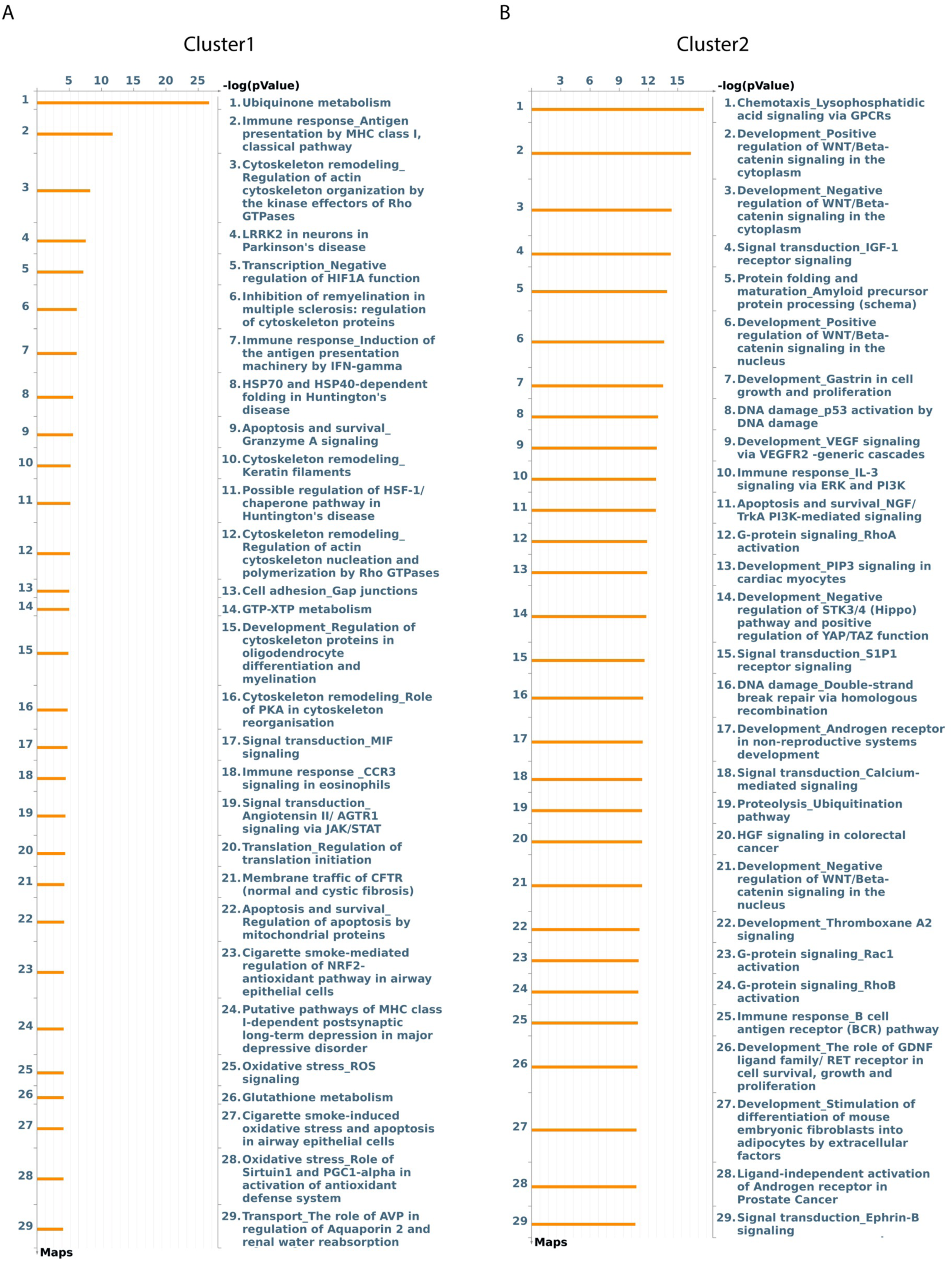
Functional classification of CUP#55 cluster markers. A) Pathway enrichment analysis of positive marker genes characterizing cluster1 and B) of cluster 2. Marker genes (P <0.05) were imported into MetaCore analytical software to determine significantly enriched pathways in each group.

**Figure S7.**
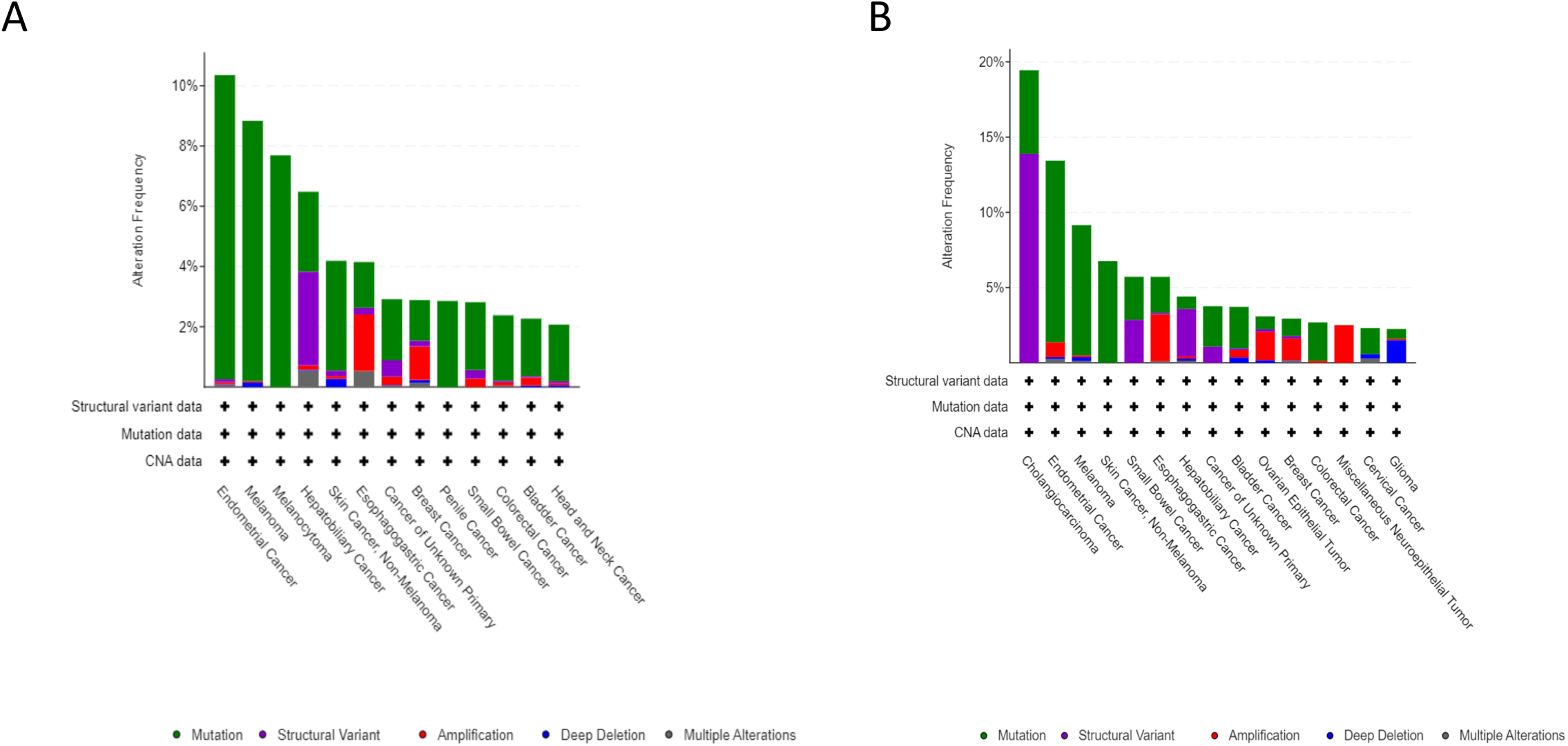
FGFR2 genomic alterations in AACR GENIE (A) and MSK-IMPACT (B) cancer types with frequency > 3%. Cancers of unknown primary were tested for genomic alterations in Zehir et al study *(16)* and AACR project GENIE *(17)*. In these studies, FGFR2 genomic alterations were detectable with a frequency of about 3-4%. The most frequent alterations were mutations, fusions, and amplifications.

**Figure S8.**
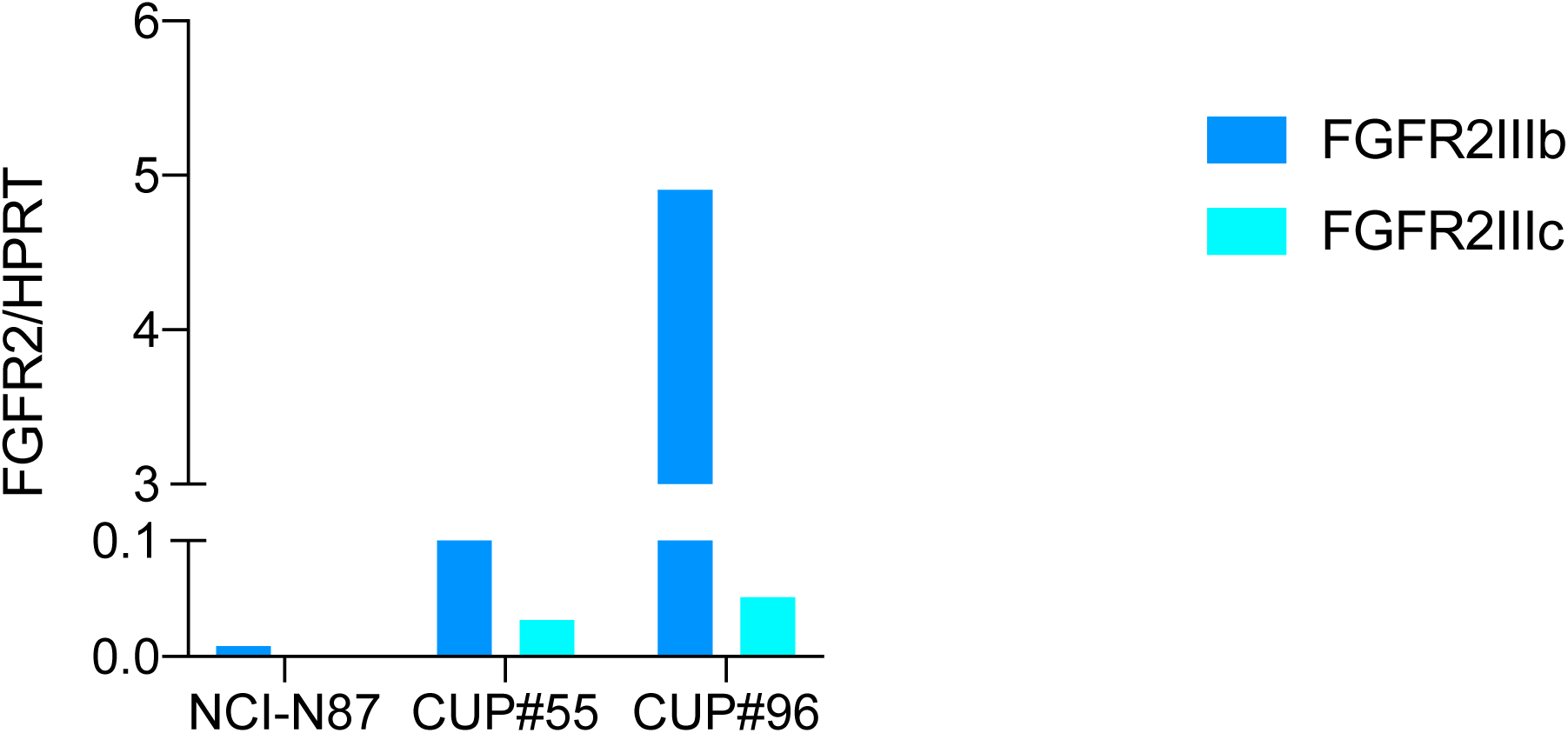
FGFR2 isoforms detection in the two cell models and a control not amplified cell line. ddPCR was used to measure the abundance of two distinct isoforms of FGFR2: FGFR2IIIb epithelial and FGFR2IIIc mesenchymal. CUP#55 and CUP#96 showed higher level of FGFR2IIIb but both expressed also the mesenchymal one suggesting an epithelial-mesenchymal transition of the cells.

**Figure S9.**
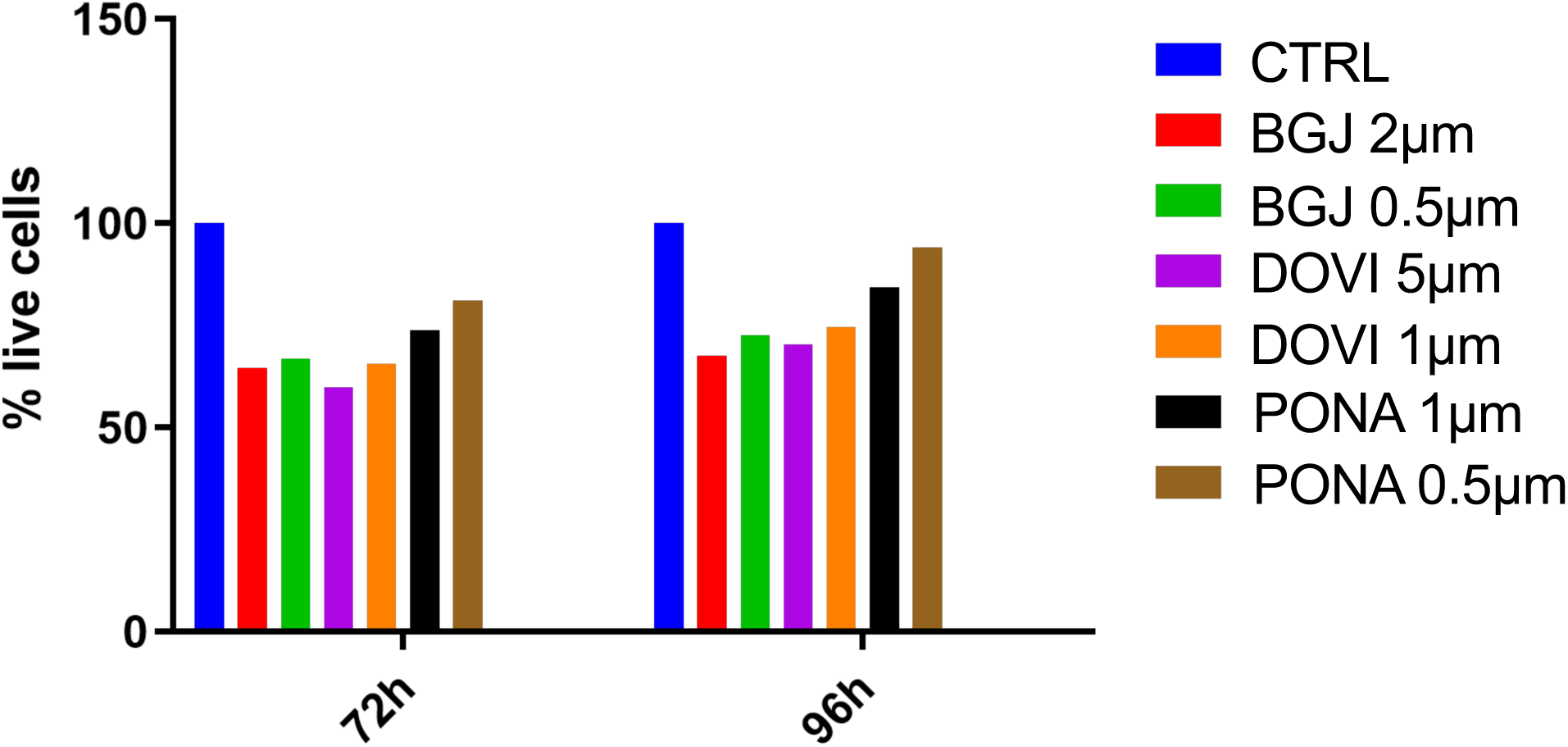
Viability assay of CUP#55 model exposed to different concentrations of FGFR2 inhibitors. Viability reduction of CUP#55 cell line after treatment with FGFR inhibitors (BGJ398, Ponatinib, Dovitinib) or cisplatin. Data were obtained using CellTiter-Glo Luminescent Cell Viability Assay (Promega).

**Figure S10.**
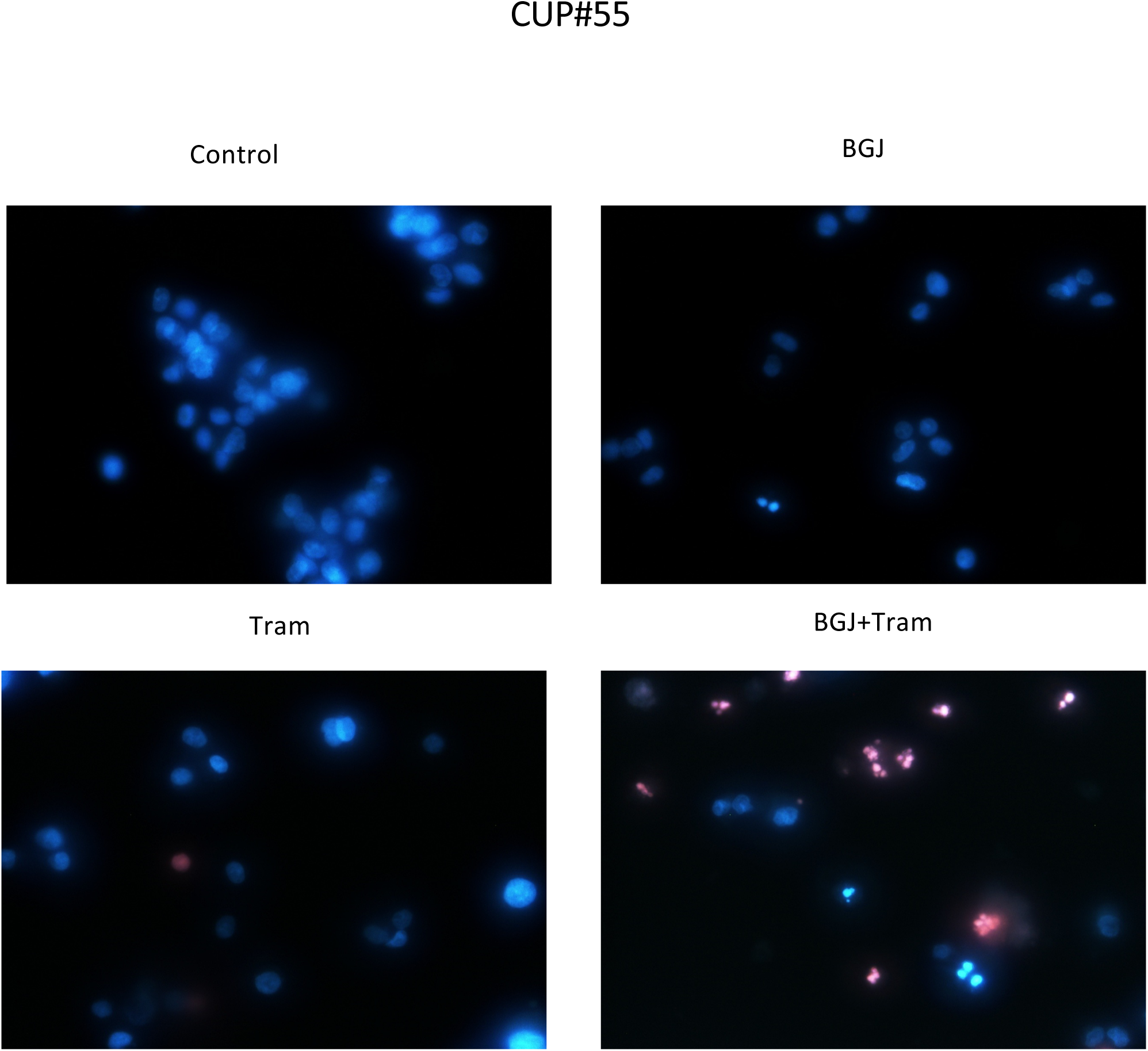
Cell death assessment upon treatment with FGFR2 and MEK inhibitors. Representative images of CUP#55 cells untreated or treated with BGJ398 1µM and/or trametinib 100 nM for 96h and analyzed by fluorescence microscopy after Hoechst 33342/PI staining (Magnification 400X).

**Table S1.**
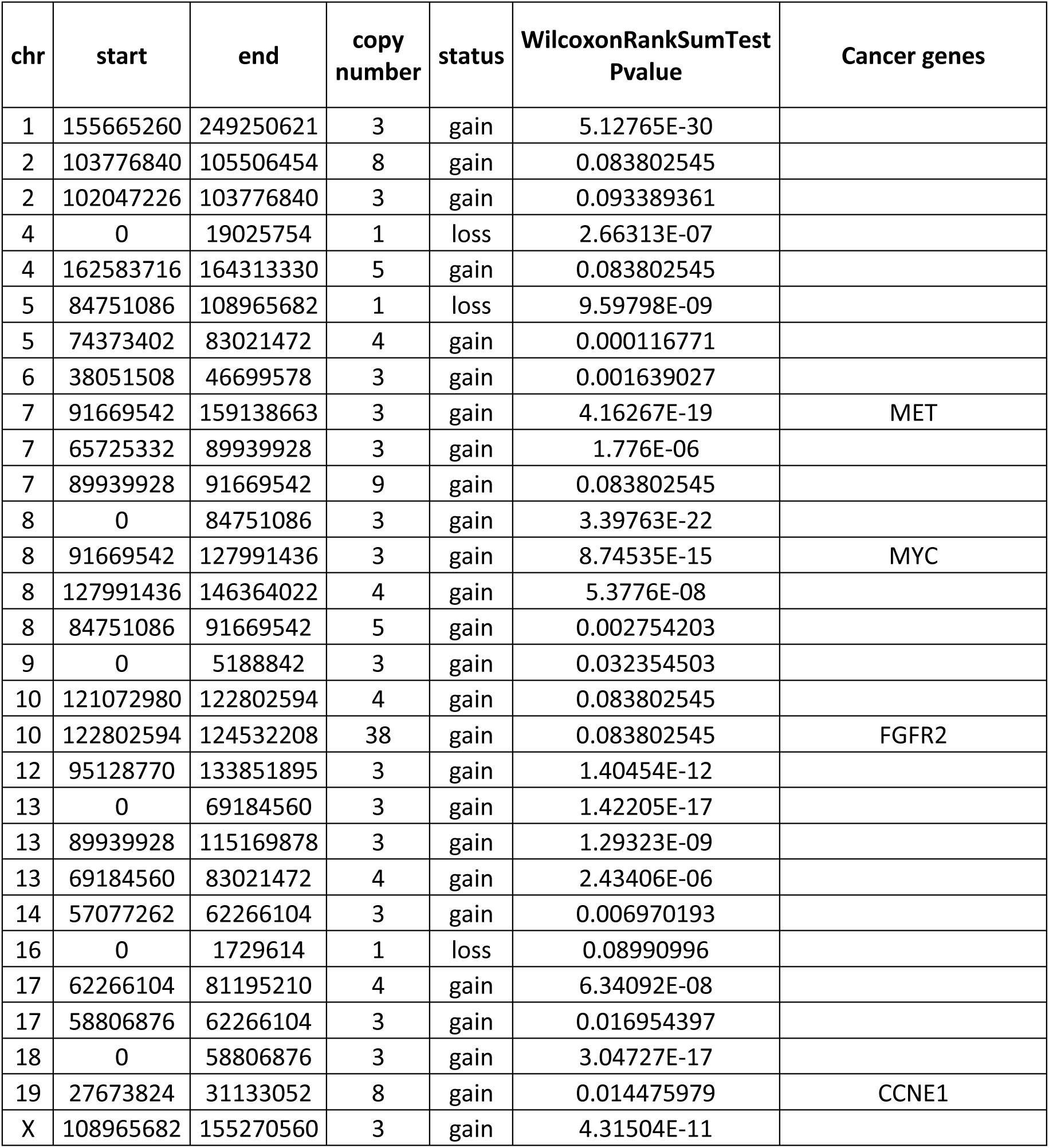
CUP#55 Copy Number Variation analysis.

| chr | start | end | copy number | status | WilcoxonRankSumTest Pvalue | Cancer genes |
| --- | --- | --- | --- | --- | --- | --- |
| 1 | 155665260 | 249250621 | 3 | gain | 5.12765E-30 |  |
| 2 | 103776840 | 105506454 | 8 | gain | 0.083802545 |  |
| 2 | 102047226 | 103776840 | 3 | gain | 0.093389361 |  |
| 4 | 0 | 19025754 | 1 | loss | 2.66313E-07 |  |
| 4 | 162583716 | 164313330 | 5 | gain | 0.083802545 |  |
| 5 | 84751086 | 108965682 | 1 | loss | 9.59798E-09 |  |
| 5 | 74373402 | 83021472 | 4 | gain | 0.000116771 |  |
| 6 | 38051508 | 46699578 | 3 | gain | 0.001639027 |  |
| 7 | 91669542 | 159138663 | 3 | gain | 4.16267E-19 | MET |
| 7 | 65725332 | 89939928 | 3 | gain | 1.776E-06 |  |
| 7 | 89939928 | 91669542 | 9 | gain | 0.083802545 |  |
| 8 | 0 | 84751086 | 3 | gain | 3.39763E-22 |  |
| 8 | 91669542 | 127991436 | 3 | gain | 8.74535E-15 | MYC |
| 8 | 127991436 | 146364022 | 4 | gain | 5.3776E-08 |  |
| 8 | 84751086 | 91669542 | 5 | gain | 0.002754203 |  |
| 9 | 0 | 5188842 | 3 | gain | 0.032354503 |  |
| 10 | 121072980 | 122802594 | 4 | gain | 0.083802545 |  |
| 10 | 122802594 | 124532208 | 38 | gain | 0.083802545 | FGFR2 |
| 12 | 95128770 | 133851895 | 3 | gain | 1.40454E-12 |  |
| 13 | 0 | 69184560 | 3 | gain | 1.42205E-17 |  |
| 13 | 89939928 | 115169878 | 3 | gain | 1.29323E-09 |  |
| 13 | 69184560 | 83021472 | 4 | gain | 2.43406E-06 |  |
| 14 | 57077262 | 62266104 | 3 | gain | 0.006970193 |  |
| 16 | 0 | 1729614 | 1 | loss | 0.08990996 |  |
| 17 | 62266104 | 81195210 | 4 | gain | 6.34092E-08 |  |
| 17 | 58806876 | 62266104 | 3 | gain | 0.016954397 |  |
| 18 | 0 | 58806876 | 3 | gain | 3.04727E-17 |  |
| 19 | 27673824 | 31133052 | 8 | gain | 0.014475979 | CCNE1 |
| X | 108965682 | 155270560 | 3 | gain | 4.31504E-11 |  |

**Table S2.**
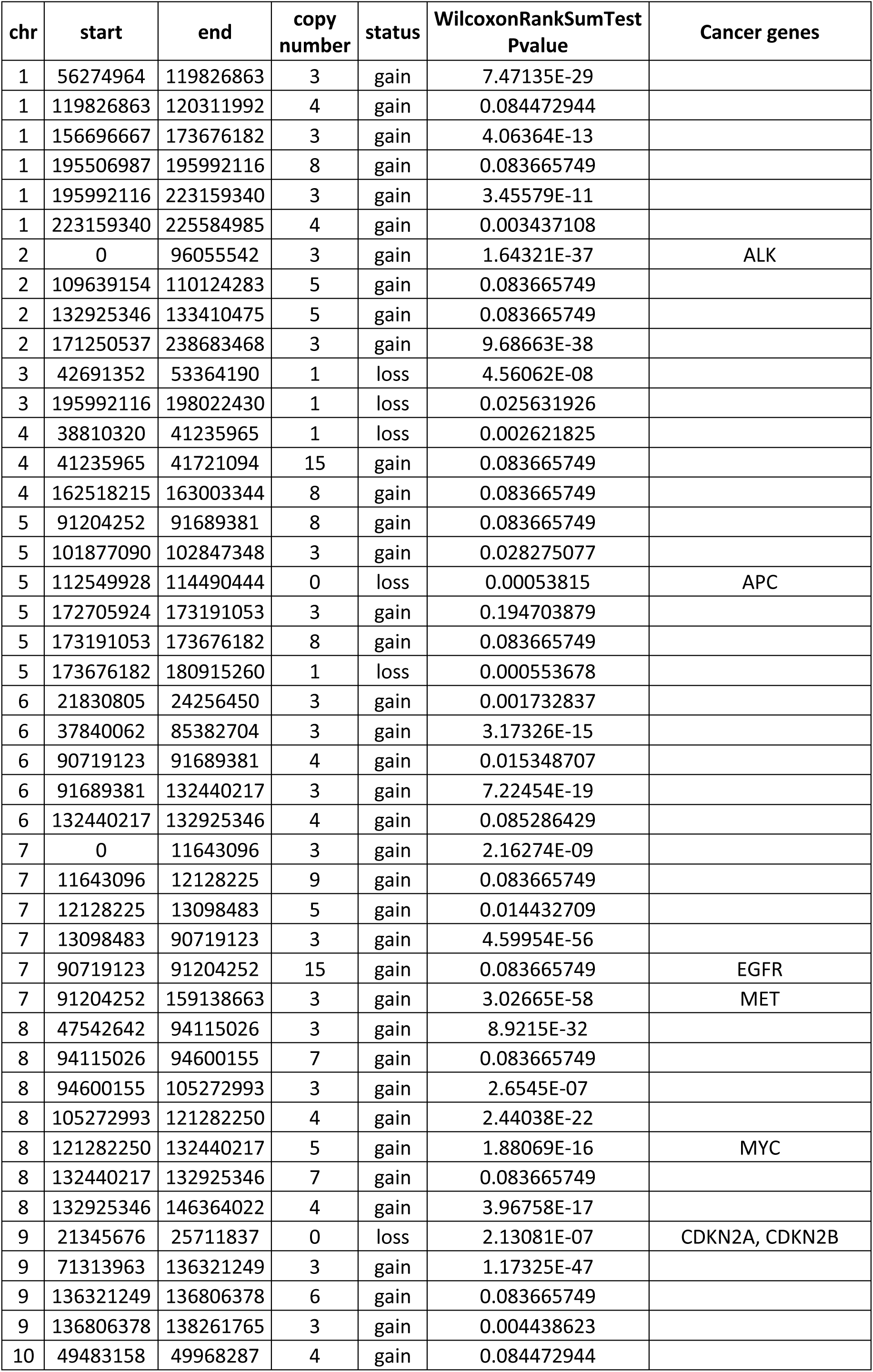
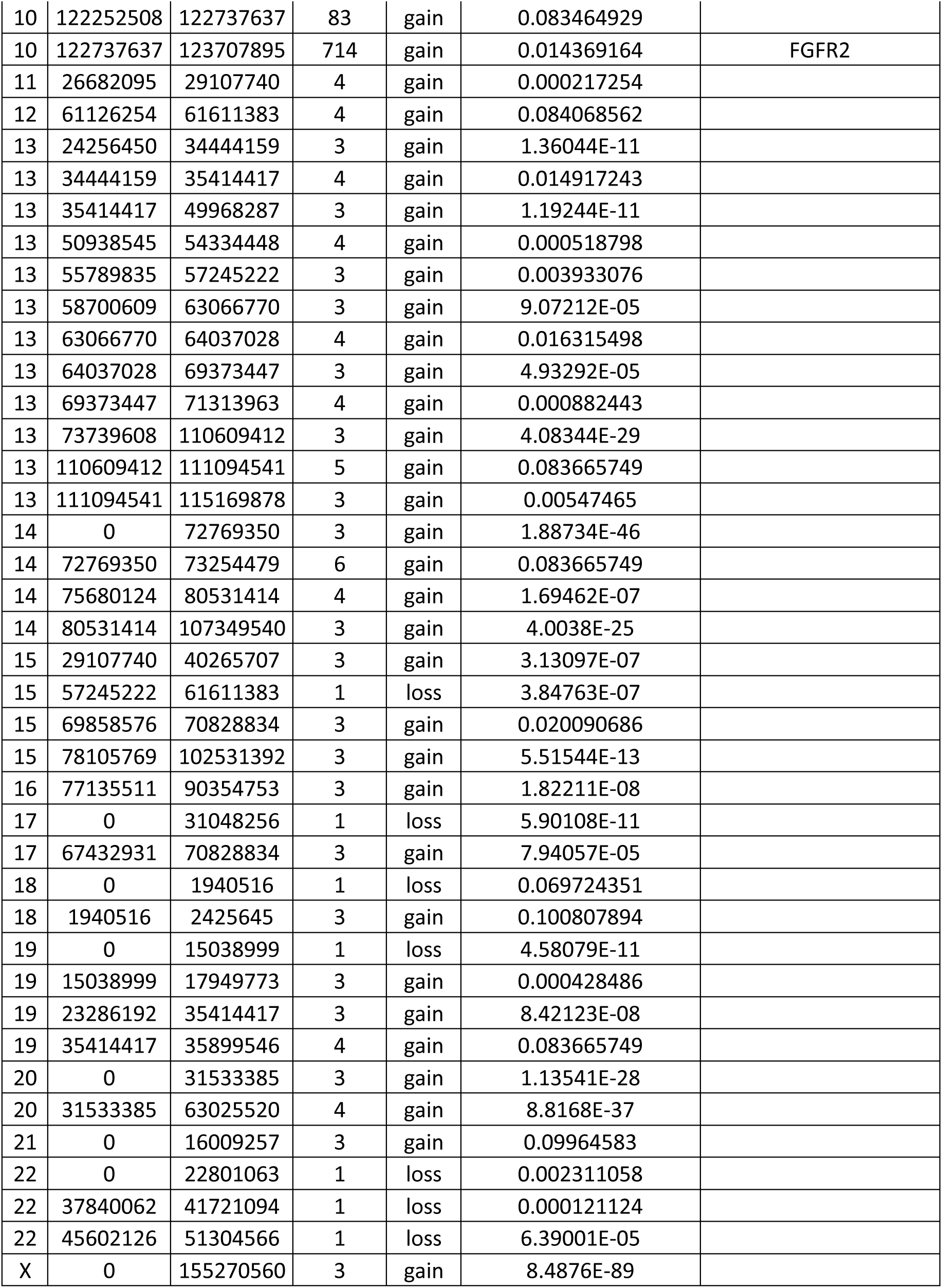
CUP#96 Copy Number Variation analysis.

